# An International Multicenter Analysis of Brain Structure across Clinical Stages of Parkinson’s Disease: The ENIGMA-Parkinson’s Study

**DOI:** 10.1101/2020.04.28.20072710

**Authors:** Max A. Laansma, Joanna K. Bright, Sarah Al-Bachari, Tim J. Anderson, Tyler Ard, Francesca Assogna, Katherine A. Baquero, Henk W. Berendse, Jamie Blair, Fernando Cendes, John C. Dalrymple-Alford, Rob M. A. de Bie, Ines Debove, Michiel F. Dirkx, Jason Druzgal, Hedley C. A. Emsley, Gäetan Garraux, Rachel P. Guimarães, Boris A. Gutman, Rick C. Helmich, Johannes C. Klein, Clare E. Mackay, Corey T. McMillan, Tracy R. Melzer, Laura M. Parkes, Fabrizio Piras, Toni L. Pitcher, Kathleen L. Poston, Mario Rango, Letícia F. Ribeiro, Cristiane S. Rocha, Christian Rummel, Lucas S. R. Santos, Reinhold Schmidt, Petra Schwingenschuh, Gianfranco Spalletta, Letizia Squarcina, Odile A. van den Heuvel, Chris Vriend, Jiun-Jie Wang, Daniel Weintraub, Roland Wiest, Clarissa L. Yasuda, Neda Jahanshad, Paul M. Thompson, Ysbrand D. van der Werf

**Affiliations:** Amsterdam UMC, Vrije Universiteit Amsterdam, Department of Anatomy & Neurosciences, Amsterdam Neuroscience, Amsterdam, The Netherlands; Imaging Genetics Center, Mark and Mary Stevens Neuroimaging and Informatics Institute, Keck School of Medicine, University of Southern California, Marina del Rey, CA, USA; Faculty of Health and Medicine, The University of Lancaster, Lancaster, UK; Division of Neuroscience and Experimental Psychology, Faculty of Biology, Medicine and Health, The University of Manchester, Manchester Academic Health Science Centre, Manchester, UK; Department of Neurology, Royal Preston Hospital, Preston, UK; Department of Medicine, University of Otago, Christchurch, Christchurch, New Zealand; USC Stevens Neuroimaging and Informatics Institute, Department of Neurology, University of Southern California, Los Angeles, CA, USA; Laboratory of Neuropsychiatry, IRCCS Santa Lucia Foundation, Rome, Italy; GIGA-CRC in vivo imaging, University of Liège, Liège, Belgium; Amsterdam UMC, University of Amsterdam, Department of Neurology, Amsterdam Neuroscience, Amsterdam, The Netherlands; Department of Medical Imaging, University of Virginia Health System, Charlottesville, VA, USA; Neuroimaging Laboratory, Department of Neurology, University of Campinas, Campinas, SP, Brazil; New Zealand Brain Research Institute, Christchurch, New Zealand; School of Psychology, Speech and Hearing, University of Canterbury, Christchurch, Canterbury, New Zealand; Brain Research New Zealand - Rangahau Roro Aotearoa, Centre of Research Excellence, New Zealand; Department of Neurology, University Hospital Bern, Inselspital, University of Bern, Bern, Switzerland; Department of Neurology and Center of Expertise for Parkinson & Movement Disorders, Donders Institute for Brain, Cognition and Behaviour, Radboud University Nijmegen Medical Centre, Nijmegen, The Netherlands; Centre for Cognitive Neuroimaging, Donders Institute for Brain, Cognition and Behaviour, Radboud University Nijmegen, Nijmegen, The Netherlands; Department of Radiology and Medical Imaging, University of Virginia, Charlottesville, VA, USA; Lancaster Medical School, Lancaster University, Preston, UK; Department of Neurology, CHU Liège, Liège, Liège, Belgium; Department of Biomedical Engineering, Illinois Institute of Technology, Chicago, IL, USA; Oxford Parkinson’s Disease Centre, Nuffield, Department of Clinical Neurosciences, Division of Clinical Neurology, University of Oxford, Oxford, UK; Department of Psychiatry, University of Oxford, Oxford, UK; University of Pennsylvania Perelman School of Medicine, Philadelphia, PA, USA; Department of Neurology & Neurological Sciences, Stanford University, Palo Alto, CA, USA; Excellence Center for Advanced MR Techniques and Parkinson’s Disease Center, Neurology unit, Fondazione IRCCS Cà Granda Maggiore Policlinico Hospital, University of Milan, Milan, Italy; Department of Medical Genetics, University of Campinas, Campinas, SP, Brazil; Support Center for Advanced Neuroimaging (SCAN), University Institute of Diagnostic and Interventional Neuroradiology, University Hospital Bern, Switzerland; Department of Neurology, Clinical Division of Neurogeriatrics, Medical University Graz, Graz, Austria; Department of Neurology, Medical University of Graz, Graz, Austria; Amsterdam UMC, Vrije Universiteit Amsterdam, Psychiatry, Amsterdam Neuroscience, Amsterdam, The Netherlands; Department of Medical Imaging and Radiological Sciences, Chang Gung University, Taoyuan City, Taiwan; Department of Diagnostic Radiology, Chang Gung Memorial Hospital, Keelung Branch Keelung City, Taiwan; Department of Psychiatry, University of Pennsylvania Perelman School of Medicine, Philadelphia, PA, USA

**Keywords:** Parkinson’s disease, structural MRI, brain, mega-analysis, ENIGMA, disease severity, clinical stages

## Abstract

**Background:** Brain structure abnormalities throughout the course of Parkinson’s disease (PD) have yet to be fully elucidated. Inconsistent findings across studies may be partly due to small sample sizes and heterogeneous analysis methods. Using a multicenter approach and harmonized analysis methods, we aimed to overcome these limitations and shed light on disease stage-specific profiles of PD pathology as suggested by *in vivo* neuroimaging.

**Methods:** Individual brain MRI and clinical data from 2,367 PD patients and 1,183 healthy controls were collected from 19 sites, deriving from 20 countries. We analyzed regional cortical thickness, cortical surface area, and subcortical volume using mixed-effect linear models. Patients were grouped according to the Hoehn & Yahr (HY) disease stages and compared to age- and sex-matched controls. Within the PD sample, we investigated associations between Montreal Cognitive Assessment (MoCA) scores and brain morphology.

**Findings:** The main analysis showed a thinner cortex in 38 of 68 regions in PD patients compared to controls (*dmax* = −0·25, *dmin* = −0·13). The bilateral putamen (left: *d* = −0·16, right: *d* = −0·16) and left amygdala (*d* = −0·15) were smaller in patients, while the left thalamus was larger (*d* = 0·17). HY staging indicated that a thinner cortex initially presents in the occipital, parietal and temporal cortex, and extends towards caudally located brain regions with increased disease severity. From HY stage 2 and onwards the bilateral putamen and amygdala were consistently smaller with larger effects denoting each increment. Finally, we found that poorer cognitive performance was associated with widespread cortical thinning as well as lower volumes of core limbic structures.

**Interpretation:** Our findings offer robust and novel imaging signatures that are specific to the disease severity stages and in line with an ongoing neurodegenerative process, highlighting the importance of such multicenter collaborations.

**Funding:** NIH Big Data to Knowledge program, ENIGMA World Aging Center, and ENIGMA Sex Differences Initiative, and other international agencies (listed in full in the Acknowledgments).

## Introduction

Parkinson’s disease (PD) is the world’s second most prevalent neurodegenerative disease. Apart from the cardinal motor symptoms, patients may suffer from cognitive, neuropsychiatric, and autonomic dysfunction.^1^ Clinical features of PD are thought to arise in part from dysfunctions of neural circuits, involving both cortical and subcortical regions.^2^ The use of neuroimaging to investigate the macroscopic brain structural changes in PD may help to understand patterns of the underlying pathology and potentially provide *in vivo* biomarkers of disease process and development.

Structural magnetic resonance imaging (MRI) of the brain allows for non-invasive assessment of cortical and subcortical morphology. Most imaging studies of PD have reported findings consistent with the atrophic process that underlies neurodegeneration, such as lower measures of subcortical volume and cortical thickness in PD compared to healthy controls.^3^ The reported atrophy patterns vary across studies in terms of location and effect size, and it is still poorly understood how disease severity relates to profiles of abnormal brain morphology.^3,4^ The discrepancies may be, in part, explained by methodological factors, including small sample sizes for individual studies, and differences in analysis methods. Heterogeneity with respect to the demographics and clinical characteristics of the patient sample, the regions of interest assessed, and the use of algorithms for segmentation and parcellation (e.g., atlas-based versus voxel- or vertex-based) may also produce differences in reported findings, which in turn makes study findings hard to compare.

Large-scale collaborations, such as the Enhancing Neuroimaging through Meta-Analysis (ENIGMA) consortium, have been initiated to overcome these limitations, by harmonizing data processing and analysis across studies, and aggregating information across multiple samples worldwide. The multicenter approach has proven effective in confining confounding factors and increasing statistical power.^5^

The ENIGMA Parkinson’s Disease (ENIGMA-PD) Working Group is an international initiative, set up to identify imaging signatures of pathology in PD, and factors that influence them. In the largest prospective cohort study on PD brain morphology to date, we report on differences in regional cortical thickness, cortical surface area, and subcortical volume derived from a comparison between PD patients and healthy control subjects, taking into account disease severity, dopamine replacement therapy, age, and sex. In addition, we investigated morphological correlates of global cognitive abilities within the PD sample. By using a mega-analytic approach and a large sample size, we aim to reveal a robust signature for the direction and magnitude of abnormalities.

## Methods

### Samples

We analyzed T1-weighted MRI scans from 19 sites, deriving from 20 countries (**Figure S1**), comprising 2,367 PD patients and 1,183 control subjects. Clinical information from the PD subjects included Hoehn and Yahr (HY) stages,^6^ duration of illness, medication status, and Montreal Cognitive Assessment (MoCA) scores.^7^ Controls were defined as individuals with no history of neurological or psychiatric illness. Individual site inclusion/exclusion criteria are provided in the supplementary material (**Table S1**). The 43 samples of Parkinson’s patients and controls provided were defined as ‘cohorts’, such that sites may contribute multiple cohorts from separate testing environments; in particular, the Parkinson’s Progression Markers Initiative (PPMI) collects imaging data across multiple clinical sites,^8^ and these were treated as independent cohorts. Disease severity was assessed using HY stages, ranging from 1 to 5; stage 1 denotes unilateral motor impairment, and stage 5 being confined to bed or wheelchair. The modified HY classification, which includes intermediate increments of 1·5 and 2·5 to complement stage 2,^9^ was used in 12 cohorts. We regrouped the cases so that stage 1·5 (N = 83) and stage 2·5 (N = 169) patients were included in the stage 2 group. Stage 4 (N = 68) and 5 (N = 17) patient groups were merged. The nearest neighbor matching procedure, featured in the MatchIt software package for R,^10^ selected an age- and sex-balanced subsample of controls for each HY stage group based on propensity score matching with replacement.

### Image Acquisition and Processing

All structural brain MRI scans were obtained with a 3-dimensional gradient-echo T1-weighted sequence. All site-specific parameters are summarized in the supplementary material (**Table S2**). Contributing sites processed their data locally using standardized ENIGMA protocols for harmonization and quality control (see http://enigma.ini.usc.edu/protocols/imaging-protocols/). Regional cortical thickness, cortical surface area, and subcortical volume metrics were extracted from the brain images using FreeSurfer 5·3. For each subject, FreeSurfer parcellated 34 cortical regions per hemisphere based on the Desikan-Killiany atlas and segmented 8 subcortical regions in each hemisphere.^11^ Regions with thickness, surface area, or volume values higher than the 75% quantile or lower than the 25% quantile were visually inspected using the original anatomical image overlaid with the parcellated/segmented image. Poorly parcellated/segmented regions were excluded from the statistical analysis, in accordance with the ENIGMA quality control protocols. All collaborators in our Working Group granted permission to share individual patient derived data (IPD), including demographic and clinical characteristics and FreeSurfer-derived measures of cortical thickness, cortical surface area, and subcortical volumes. All sites provided anonymized data with ethical approval from their local ethics committees and institutional review boards.

### Mega-Analysis of Cortical and Subcortical Properties

Between-group differences were assessed using multiple linear mixed-effects regressions on the pooled means of cortical thickness (in mm), surface area (in mm^2^), and subcortical volume (in mm^3^). Primary model 1a included age, sex, and intracranial volume (ICV) as fixed effects and cohort as a random effect. Adjusted primary models omitted ICV (model 1b), age and ICV (model 1c), age and sex (model 2a), or age, sex, and ICV (model 2b). Secondary models 3a,b included an interaction term - sex-by-diagnosis - thereby extending model 1a,b, respectively (**Figure 1**).

**Figure 1.**
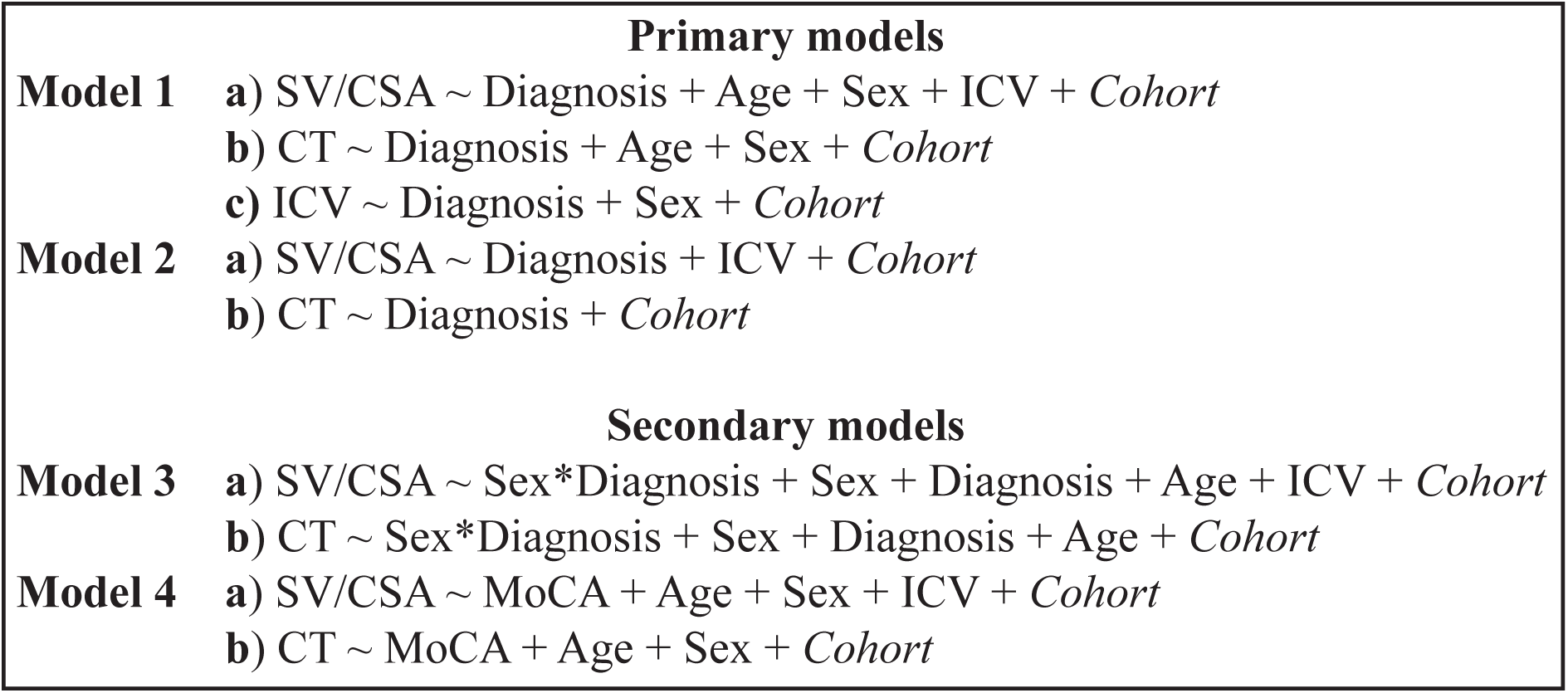
Summary of the models used. The statistical models included cortical thickness, cortical surface area, and subcortical volume as dependent variables, Diagnosis, Age, Sex, and ICV as independent fixed variables, and Cohort as a random variable. The analyses were performed for 68 cortical and 16 subcortical ROIs. Model 1 also included ICV for the subcortical analysis. Abbreviations: ROI, region of interest; CT, cortical thickness; CSA, cortical surface area; SV, subcortical volume; ICV, intracranial volume; MoCA, Montreal Cognitive Assessment.

The main analysis examined differences between all patients and controls using model 1a for subcortical volume and cortical surface area, model 1b for cortical thickness, and model 1c for ICV; omitting ICV in the thickness model is appropriate to previous research on nuisance factors.^12^ Differences between patients grouped by HY stage and controls were assessed using model 2a for subcortical volume and cortical surface area and model 2b for cortical thickness. The medication analysis, which compared patients receiving dopaminergic treatment (i.e., medicated) versus treatment-naive patients (i.e., unmedicated) within HY stage 1 and 2, was performed using model 2a for subcortical volume and cortical surface area and model 2b for cortical thickness. We investigated the interaction between sex and diagnosis at each HY stage using model 3a for subcortical volume and cortical surface area and model 3b for cortical thickness. The partial Cohen’s *d* was estimated from the *t*-statistic to quantify the effect size of the differences. The percentage difference of patients from controls was calculated using the least square group means.

In addition, we used a linear mixed-effects regression model to examine within-group associations between cognitive ability and morphometry, incorporating a variant of model 1 for subcortical volume (secondary model 4a) and cortical surface area, and model 2 for cortical thickness (secondary model 4b). The *t*-statistic was reported as the effect size. To determine how representative the MoCA subgroup was for the complete PD sample, we performed a differential analysis between the PD group with available MoCA scores and the control group (supplementary material).

Significant results that passed the conservative Bonferroni correction for multiple comparisons were reported; the significance level for each statistical model was determined by dividing the *p* value 0·05 by the number of ROIs.

## Results

### Sample

Data flow for each analysis is depicted in **Figure 2**. Data from a total of 2,367 patients and 1,183 controls were available for the analyses (**Table 1**).

**Figure 2.**
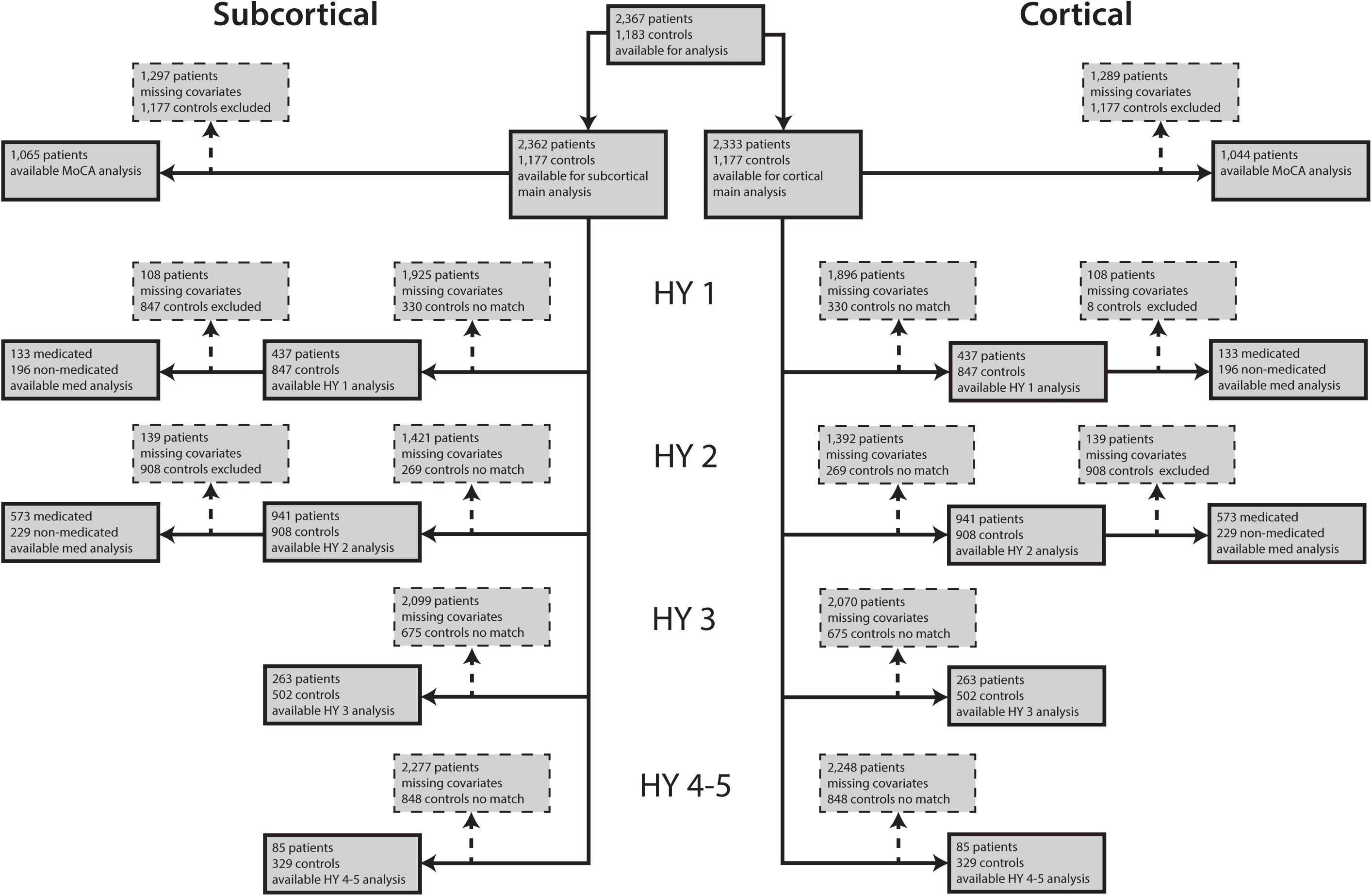
Flowchart of data inclusion. Schematic overview of derived subcortical and cortical samples for each analysis. Abbreviations: HY, Hoehn & Yahr; med, medication; MoCA, Montreal Cognitive Assessment.

**Table 1:** Analysis sample characteristics.

| Site | Cohort | N |  | Mean age, years (SD) |  | % Female |  | DURILL,<br>years (SD) | MoCA (SD) |
| --- | --- | --- | --- | --- | --- | --- | --- | --- | --- |
|  |  | HC | PD | HC | PD | HC | PD | PD | PD |
| Amsterdam | Amsterdam I | 44 | 138 | 56.5 (9.48) | 63.1 (10.81) | 39 | 38 | 2.1 (3.33) | NA |
|  | Amsterdam II (Cogtips) | 0 | 61 | NA | 62.5 (7.08) | NA | 39 | 5.3 (3.54) | 26.3 (2.15) |
| Bern | BE I | 23 | 52 | 54.1 (9.78) | 62.9 (10.38) | 30 | 52 | 12.4 (4.29) | 23 (5.66) |
|  | BE II | 30 | 3 | 68.2 (4.45) | 59.7 (6.66) | 70 | 67 | 11.3 (7.57) | NA |
| Campinas | UNICAMP | 138 | 110 | 58.9 (7.91) | 60.0 (10.36) | 63 | 34 | 6.9 (6.22) | NA |
| Chang Gung | CGU | 223 | 327 | 61.0 (7.25) | 60.1 (9.63) | 54 | 43 | 8.6 (6.32) | NA |
| Charlottesville | UVA I | 0 | 116 | NA | 63.7 (8.52) | NA | 28 | 9.7 (5.09) | 24.9 (3.44) |
|  | UVA II | 0 | 37 | NA | 62.4 (9.59) | NA | 14 | 8.7 (3.64) | 24.3 (3.45) |
|  | UVA III | 0 | 24 | NA | 70.8 (6.77) | NA | 29 | 7.7 (3.23) | 23.4 (4.60) |
| Christchurch | PDNZ | 39 | 209 | 67.5 (8.14) | 69.4 (7.77) | 33 | 26 | 5.7 (5.57) | 23.6 (4.18) |
| Donders | Donders | 23 | 59 | 62.7 (10.29) | 60.8 (10.07) | 48 | 44 | 4.4 (3.79) | NA |
| Graz | PROMOVE/ASPS | 124 | 100 | 63.4 (10.07) | 63.2 (10.15) | 27 | 29 | 4.7 (4.77) | NA |
|  | PROMOVE/ASPS | 0 | 23 | NA | 64.0 (9.90) | NA | 22 | 4.0 (5.69) | NA |
| Liege | Liege I | 33 | 30 | 65.8 (4.29) | 65.9 (6.61) | 45 | 37 | 7.2 (5.32) | NA |
|  | Liege II | 43 | 45 | 64.8 (8.33) | 66.9 (8.24) | 49 | 44 | 6 (3.93) | NA |
| Milan | Milan | 10 | 44 | 53.3 (10.53) | 57.8 (7.71) | 70 | 32 | 11.4 (3.38) | NA |
| NW-England | NW-England I | 22 | 32 | 70.6 (7.92) | 69.9 (8.58) | 45 | 19 | 6.8 (4.42) | 24.8 (4.25) |
|  | NW-England II | 13 | 14 | 64.6 (3.99) | 65.0 (5.67) | 38 | 29 | 9.2 (6.02) | 26.3 (2.93) |
| NEUROCON | NEUROCON | 15 | 27 | 66.7 (11.74) | 68.7 (10.55) | 80 | 37 | NA | NA |
| ON Japan | ON Japan | 15 | 30 | 63.3 (5.25) | 67.6 (6.81) | 53 | 57 | NA | NA |
| Oxford | Oxford Discovery | 57 | 115 | 65.6 (8.67) | 63.9 (10.05) | 39 | 36 | 2.3 (1.58) | 26.5 (2.73) |
| Pennsylvania | UDALL/U19 | 11 | 122 | 70.1 (5.86) | 66.4 (7.87) | 55 | 33 | 8 (5.34) | 24.7 (5.04) |
| PPMI | PPMI 1-21 | 163 | 347 | 60.2 (11.64) | 61.7 (9.65) | 36 | 35 | 0.6 (0.56) | 27.1 (2.26) |
| Rome SLF | Rome SLF | 125 | 239 | 36.6 (10.63) | 62.7 (10.25) | 41 | 37 | 4.9 (4.14) | NA |
| Stanford | Stanford | 12 | 44 | 66.3 (6.74) | 68.6 (8.49) | 75 | 50 | 5.6 (3.44) | 23.9 (5.19) |
| Tao Wu | Tao Wu | 20 | 19 | 64.8 (5.58) | 65.0 (4.45) | 40 | 47 | 5.3 (4.00) | NA |
| **Total** |  | **1183** | **2367** | **59.4 (12.33)** | **63.4 (9.78)** | **46** | **36** | **5.5 (5.44)** | **25.6 (3.70)** |

### Cortex

Main cortical analysis showed a significantly thinner cortex in PD patients compared to controls in 20 of the 34 left hemisphere regions (*dmax* = −0·25, −1·79%; *dmin* = −0·13, −0·78%) and 18 of the 34 right hemisphere regions (*dmax* = −0·25, −1·89%; *dmin* = −0·13, −0·88%; **Figure 3a** and **Table 2a**). The effects appeared symmetrical in 16 regions. Surface areas of the left frontal pole (*d* = −0·20, −3·07%) and lateral occipital cortex (*d* = −0·13, −1·45%) were significantly smaller in PD patients (**Figure 3b** and **Table 2b**). No significant differences in surface area were found in the right hemisphere.

**Figure 3.**
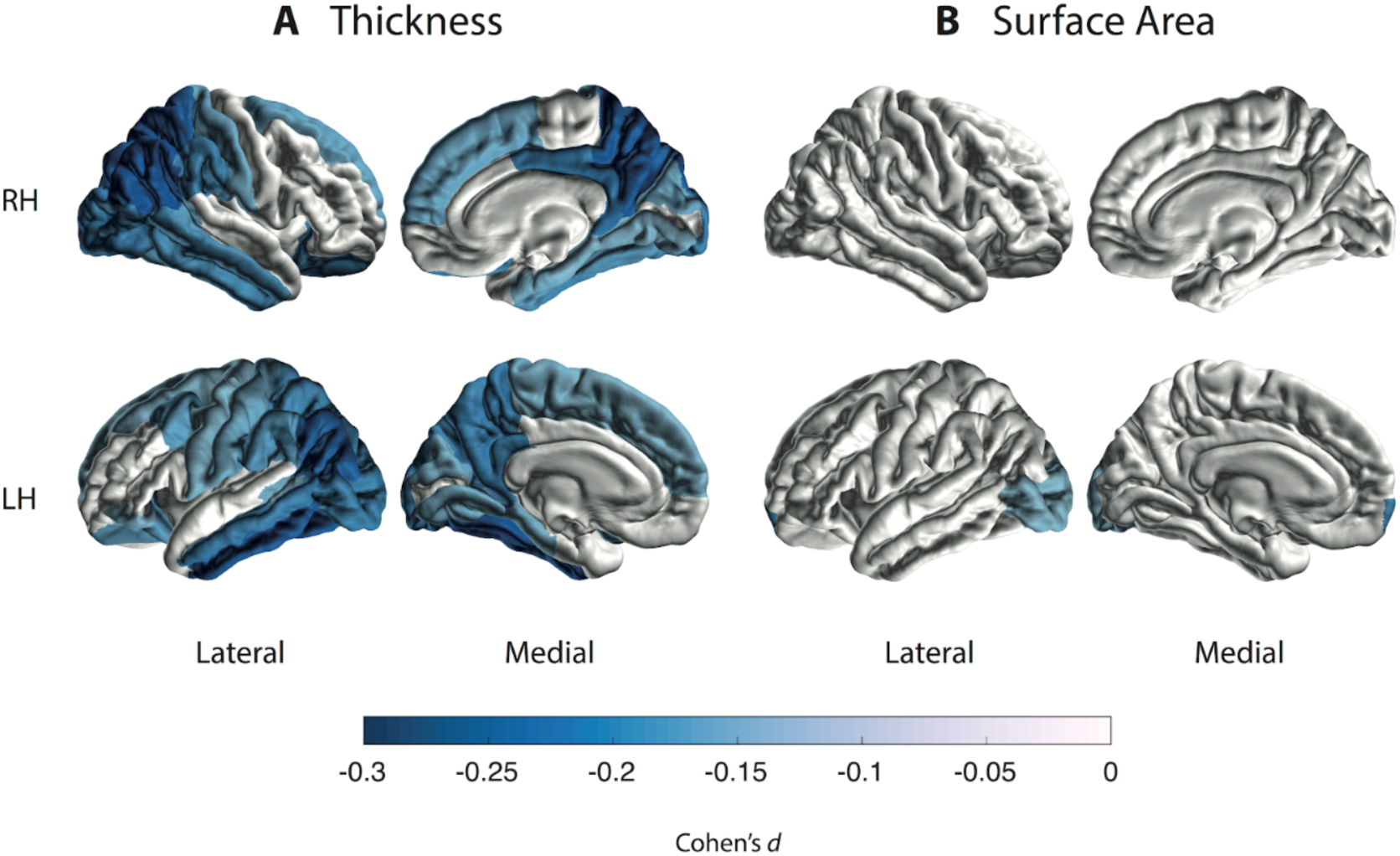
Cortical thickness and surface area group differences for PD patients vs controls. Cohen’s *d* effect size estimates are shown, for differences in (**A**) cortical thickness and (**B**) cortical surface area. A negative *d*-value indicates smaller measurements in PD patients. Cortical regions with *P*-values < 7·35 × 10^−4^ (i.e., 0·05/68 ROIs) are depicted in the heatmap colors. Abbreviations: RH, right hemisphere; LH, left hemisphere; ROI, region of interest.

**Table 2a:**
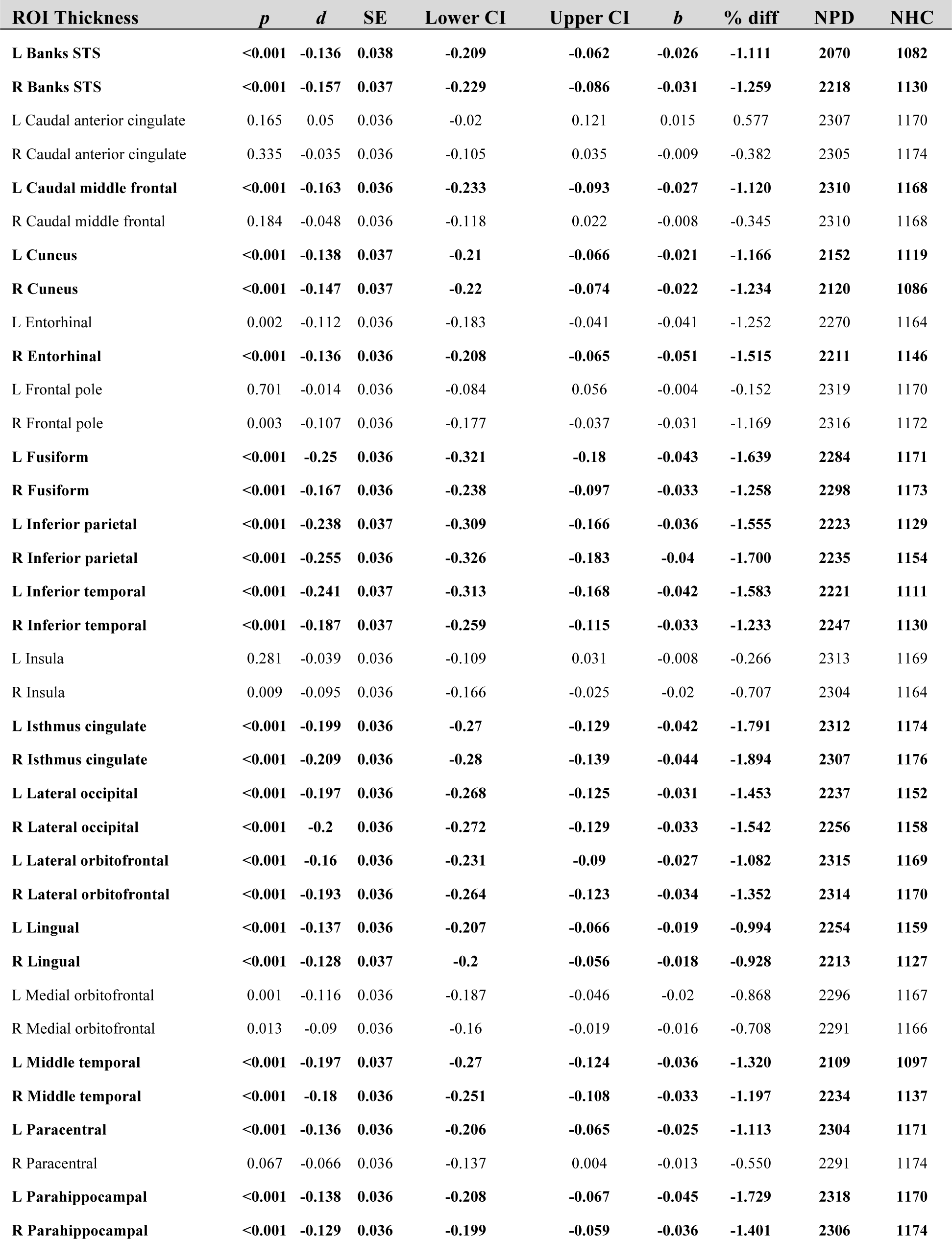
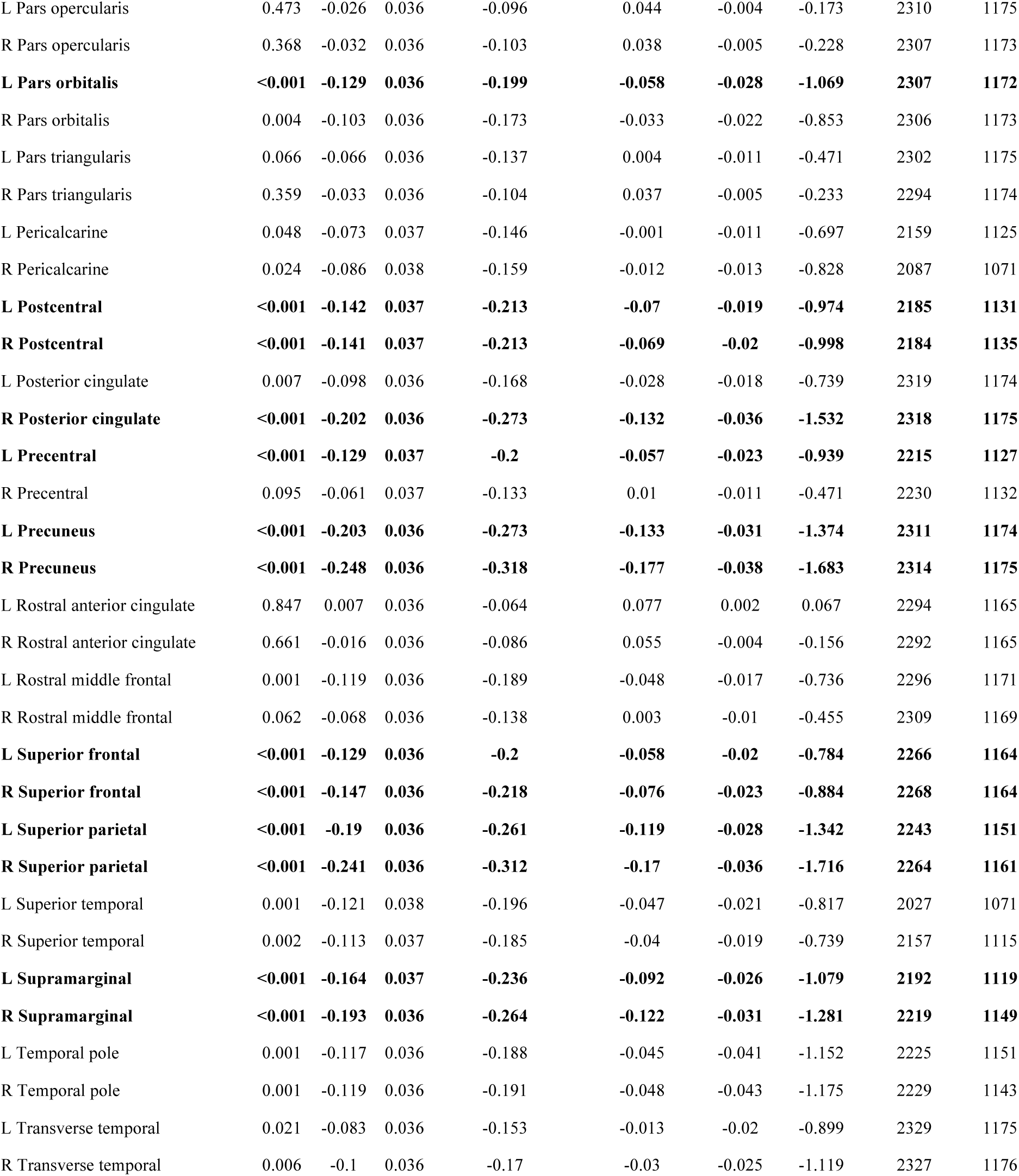
Cortical thickness in PD vs controls.

| ROI Thickness | *p* | *d* | SE | Lower CI | Upper CI | *b* | % diff | NPD | NHC |
| --- | --- | --- | --- | --- | --- | --- | --- | --- | --- |
| **L Banks STS** | **<0.001** | **-0.136** | **0.038** | **-0.209** | **-0.062** | **-0.026** | **-1.111** | **2070** | **1082** |
| **R Banks STS** | **<0.001** | **-0.157** | **0.037** | **-0.229** | **-0.086** | **-0.031** | **-1.259** | **2218** | **1130** |
| L Caudal anterior cingulate | 0.165 | 0.05 | 0.036 | -0.02 | 0.121 | 0.015 | 0.577 | 2307 | 1170 |
| R Caudal anterior cingulate | 0.335 | -0.035 | 0.036 | -0.105 | 0.035 | -0.009 | -0.382 | 2305 | 1174 |
| **L Caudal middle frontal** | **<0.001** | **-0.163** | **0.036** | **-0.233** | **-0.093** | **-0.027** | **-1.120** | **2310** | **1168** |
| R Caudal middle frontal | 0.184 | -0.048 | 0.036 | -0.118 | 0.022 | -0.008 | -0.345 | 2310 | 1168 |
| **L Cuneus** | **<0.001** | **-0.138** | **0.037** | **-0.21** | **-0.066** | **-0.021** | **-1.166** | **2152** | **1119** |
| **R Cuneus** | **<0.001** | **-0.147** | **0.037** | **-0.22** | **-0.074** | **-0.022** | **-1.234** | **2120** | **1086** |
| L Entorhinal | 0.002 | -0.112 | 0.036 | -0.183 | -0.041 | -0.041 | -1.252 | 2270 | 1164 |
| **R Entorhinal** | **<0.001** | **-0.136** | **0.036** | **-0.208** | **-0.065** | **-0.051** | **-1.515** | **2211** | **1146** |
| L Frontal pole | 0.701 | -0.014 | 0.036 | -0.084 | 0.056 | -0.004 | -0.152 | 2319 | 1170 |
| R Frontal pole | 0.003 | -0.107 | 0.036 | -0.177 | -0.037 | -0.031 | -1.169 | 2316 | 1172 |
| **L Fusiform** | **<0.001** | **-0.25** | **0.036** | **-0.321** | **-0.18** | **-0.043** | **-1.639** | **2284** | **1171** |
| **R Fusiform** | **<0.001** | **-0.167** | **0.036** | **-0.238** | **-0.097** | **-0.033** | **-1.258** | **2298** | **1173** |
| **L Inferior parietal** | **<0.001** | **-0.238** | **0.037** | **-0.309** | **-0.166** | **-0.036** | **-1.555** | **2223** | **1129** |
| **R Inferior parietal** | **<0.001** | **-0.255** | **0.036** | **-0.326** | **-0.183** | **-0.04** | **-1.700** | **2235** | **1154** |
| **L Inferior temporal** | **<0.001** | **-0.241** | **0.037** | **-0.313** | **-0.168** | **-0.042** | **-1.583** | **2221** | **1111** |
| **R Inferior temporal** | **<0.001** | **-0.187** | **0.037** | **-0.259** | **-0.115** | **-0.033** | **-1.233** | **2247** | **1130** |
| L Insula | 0.281 | -0.039 | 0.036 | -0.109 | 0.031 | -0.008 | -0.266 | 2313 | 1169 |
| R Insula | 0.009 | -0.095 | 0.036 | -0.166 | -0.025 | -0.02 | -0.707 | 2304 | 1164 |
| **L Isthmus cingulate** | **<0.001** | **-0.199** | **0.036** | **-0.27** | **-0.129** | **-0.042** | **-1.791** | **2312** | **1174** |
| **R Isthmus cingulate** | **<0.001** | **-0.209** | **0.036** | **-0.28** | **-0.139** | **-0.044** | **-1.894** | **2307** | **1176** |
| **L Lateral occipital** | **<0.001** | **-0.197** | **0.036** | **-0.268** | **-0.125** | **-0.031** | **-1.453** | **2237** | **1152** |
| **R Lateral occipital** | **<0.001** | **-0.2** | **0.036** | **-0.272** | **-0.129** | **-0.033** | **-1.542** | **2256** | **1158** |
| **L Lateral orbitofrontal** | **<0.001** | **-0.16** | **0.036** | **-0.231** | **-0.09** | **-0.027** | **-1.082** | **2315** | **1169** |
| **R Lateral orbitofrontal** | **<0.001** | **-0.193** | **0.036** | **-0.264** | **-0.123** | **-0.034** | **-1.352** | **2314** | **1170** |
| **L Lingual** | **<0.001** | **-0.137** | **0.036** | **-0.207** | **-0.066** | **-0.019** | **-0.994** | **2254** | **1159** |
| **R Lingual** | **<0.001** | **-0.128** | **0.037** | **-0.2** | **-0.056** | **-0.018** | **-0.928** | **2213** | **1127** |
| L Medial orbitofrontal | 0.001 | -0.116 | 0.036 | -0.187 | -0.046 | -0.02 | -0.868 | 2296 | 1167 |
| R Medial orbitofrontal | 0.013 | -0.09 | 0.036 | -0.16 | -0.019 | -0.016 | -0.708 | 2291 | 1166 |
| **L Middle temporal** | **<0.001** | **-0.197** | **0.037** | **-0.27** | **-0.124** | **-0.036** | **-1.320** | **2109** | **1097** |
| **R Middle temporal** | **<0.001** | **-0.18** | **0.036** | **-0.251** | **-0.108** | **-0.033** | **-1.197** | **2234** | **1137** |
| **L Paracentral** | **<0.001** | **-0.136** | **0.036** | **-0.206** | **-0.065** | **-0.025** | **-1.113** | **2304** | **1171** |
| R Paracentral | 0.067 | -0.066 | 0.036 | -0.137 | 0.004 | -0.013 | -0.550 | 2291 | 1174 |
| **L Parahippocampal** | **<0.001** | **-0.138** | **0.036** | **-0.208** | **-0.067** | **-0.045** | **-1.729** | **2318** | **1170** |
| **R Parahippocampal** | **<0.001** | **-0.129** | **0.036** | **-0.199** | **-0.059** | **-0.036** | **-1.401** | **2306** | **1174** |
| L Pars opercularis | 0.473 | -0.026 | 0.036 | -0.096 | 0.044 | -0.004 | -0.173 | 2310 | 1175 |
| R Pars opercularis | 0.368 | -0.032 | 0.036 | -0.103 | 0.038 | -0.005 | -0.228 | 2307 | 1173 |
| **L Pars orbitalis** | **<0.001** | **-0.129** | **0.036** | **-0.199** | **-0.058** | **-0.028** | **-1.069** | **2307** | **1172** |
| R Pars orbitalis | 0.004 | -0.103 | 0.036 | -0.173 | -0.033 | -0.022 | -0.853 | 2306 | 1173 |
| L Pars triangularis | 0.066 | -0.066 | 0.036 | -0.137 | 0.004 | -0.011 | -0.471 | 2302 | 1175 |
| R Pars triangularis | 0.359 | -0.033 | 0.036 | -0.104 | 0.037 | -0.005 | -0.233 | 2294 | 1174 |
| L Pericalcarine | 0.048 | -0.073 | 0.037 | -0.146 | -0.001 | -0.011 | -0.697 | 2159 | 1125 |
| R Pericalcarine | 0.024 | -0.086 | 0.038 | -0.159 | -0.012 | -0.013 | -0.828 | 2087 | 1071 |
| **L Postcentral** | **<0.001** | **-0.142** | **0.037** | **-0.213** | **-0.07** | **-0.019** | **-0.974** | **2185** | **1131** |
| **R Postcentral** | **<0.001** | **-0.141** | **0.037** | **-0.213** | **-0.069** | **-0.02** | **-0.998** | **2184** | **1135** |
| L Posterior cingulate | 0.007 | -0.098 | 0.036 | -0.168 | -0.028 | -0.018 | -0.739 | 2319 | 1174 |
| **R Posterior cingulate** | **<0.001** | **-0.202** | **0.036** | **-0.273** | **-0.132** | **-0.036** | **-1.532** | **2318** | **1175** |
| **L Precentral** | **<0.001** | **-0.129** | **0.037** | **-0.2** | **-0.057** | **-0.023** | **-0.939** | **2215** | **1127** |
| R Precentral | 0.095 | -0.061 | 0.037 | -0.133 | 0.01 | -0.011 | -0.471 | 2230 | 1132 |
| **L Precuneus** | **<0.001** | **-0.203** | **0.036** | **-0.273** | **-0.133** | **-0.031** | **-1.374** | **2311** | **1174** |
| **R Precuneus** | **<0.001** | **-0.248** | **0.036** | **-0.318** | **-0.177** | **-0.038** | **-1.683** | **2314** | **1175** |
| L Rostral anterior cingulate | 0.847 | 0.007 | 0.036 | -0.064 | 0.077 | 0.002 | 0.067 | 2294 | 1165 |
| R Rostral anterior cingulate | 0.661 | -0.016 | 0.036 | -0.086 | 0.055 | -0.004 | -0.156 | 2292 | 1165 |
| L Rostral middle frontal | 0.001 | -0.119 | 0.036 | -0.189 | -0.048 | -0.017 | -0.736 | 2296 | 1171 |
| R Rostral middle frontal | 0.062 | -0.068 | 0.036 | -0.138 | 0.003 | -0.01 | -0.455 | 2309 | 1169 |
| **L Superior frontal** | **<0.001** | **-0.129** | **0.036** | **-0.2** | **-0.058** | **-0.02** | **-0.784** | **2266** | **1164** |
| **R Superior frontal** | **<0.001** | **-0.147** | **0.036** | **-0.218** | **-0.076** | **-0.023** | **-0.884** | **2268** | **1164** |
| **L Superior parietal** | **<0.001** | **-0.19** | **0.036** | **-0.261** | **-0.119** | **-0.028** | **-1.342** | **2243** | **1151** |
| **R Superior parietal** | **<0.001** | **-0.241** | **0.036** | **-0.312** | **-0.17** | **-0.036** | **-1.716** | **2264** | **1161** |
| L Superior temporal | 0.001 | -0.121 | 0.038 | -0.196 | -0.047 | -0.021 | -0.817 | 2027 | 1071 |
| R Superior temporal | 0.002 | -0.113 | 0.037 | -0.185 | -0.04 | -0.019 | -0.739 | 2157 | 1115 |
| **L Supramarginal** | **<0.001** | **-0.164** | **0.037** | **-0.236** | **-0.092** | **-0.026** | **-1.079** | **2192** | **1119** |
| **R Supramarginal** | **<0.001** | **-0.193** | **0.036** | **-0.264** | **-0.122** | **-0.031** | **-1.281** | **2219** | **1149** |
| L Temporal pole | 0.001 | -0.117 | 0.036 | -0.188 | -0.045 | -0.041 | -1.152 | 2225 | 1151 |
| R Temporal pole | 0.001 | -0.119 | 0.036 | -0.191 | -0.048 | -0.043 | -1.175 | 2229 | 1143 |
| L Transverse temporal | 0.021 | -0.083 | 0.036 | -0.153 | -0.013 | -0.02 | -0.899 | 2329 | 1175 |
| R Transverse temporal | 0.006 | -0.1 | 0.036 | -0.17 | -0.03 | -0.025 | -1.119 | 2327 | 1176 |

**Table 2b:** Cortical surface in PD vs controls.

| ROI Surface Area | *p* | *d* | SE | Lower CI | Upper CI | *b* | % diff | NPD | NHC |
| --- | --- | --- | --- | --- | --- | --- | --- | --- | --- |
| L Banks STS | 0.939 | -0.003 | 0.038 | -0.077 | 0.071 | -0.438 | -0.044 | 2068 | 1070 |
| R Banks STS | 0.277 | 0.04 | 0.037 | -0.032 | 0.112 | 5.292 | 0.582 | 2219 | 1132 |
| L Caudal anterior cingulate | 0.13 | 0.055 | 0.036 | -0.016 | 0.125 | 7.334 | 1.177 | 2306 | 1169 |
| R Caudal anterior cingulate | 0.535 | 0.022 | 0.036 | -0.048 | 0.093 | 3.45 | 0.460 | 2301 | 1173 |
| L Caudal middle frontal | 0.267 | 0.04 | 0.036 | -0.03 | 0.111 | 13.214 | 0.597 | 2307 | 1166 |
| R Caudal middle frontal | 0.961 | -0.002 | 0.036 | -0.072 | 0.069 | -0.592 | -0.029 | 2311 | 1167 |
| L Cuneus | 0.002 | -0.116 | 0.037 | -0.189 | -0.044 | -23.415 | -1.615 | 2152 | 1111 |
| R Cuneus | 0.002 | -0.12 | 0.038 | -0.193 | -0.046 | -23.869 | -1.591 | 2116 | 1064 |
| L Entorhinal | 0.437 | -0.028 | 0.036 | -0.099 | 0.042 | -2.474 | -0.598 | 2270 | 1163 |
| R Entorhinal | 0.142 | -0.054 | 0.036 | -0.125 | 0.017 | -4.291 | -1.199 | 2214 | 1144 |
| **L Frontal pole** | **<0.001** | **-0.195** | **0.036** | **-0.266** | **-0.125** | **-6.495** | **-3.066** | **2317** | **1170** |
| R Frontal pole | 0.187 | -0.048 | 0.036 | -0.118 | 0.023 | -2.024 | -0.730 | 2318 | 1172 |
| L Fusiform | 0.24 | -0.043 | 0.036 | -0.113 | 0.028 | -15.182 | -0.488 | 2284 | 1171 |
| R Fusiform | 0.21 | -0.045 | 0.036 | -0.116 | 0.025 | -16.041 | -0.527 | 2298 | 1172 |
| L Inferior parietal | 0.412 | -0.03 | 0.037 | -0.102 | 0.041 | -16.386 | -0.369 | 2224 | 1128 |
| R Inferior parietal | 0.802 | 0.009 | 0.036 | -0.062 | 0.08 | 5.719 | 0.109 | 2239 | 1154 |
| L Inferior temporal | 0.091 | -0.063 | 0.037 | -0.135 | 0.009 | -26.378 | -0.821 | 2224 | 1108 |
| R Inferior temporal | 0.161 | -0.051 | 0.036 | -0.123 | 0.02 | -19.915 | -0.652 | 2249 | 1129 |
| L Insula | 0.593 | 0.019 | 0.036 | -0.051 | 0.09 | 4.267 | 0.194 | 2312 | 1161 |
| R Insula | 0.964 | -0.002 | 0.036 | -0.072 | 0.069 | -0.42 | -0.019 | 2304 | 1157 |
| L Isthmus cingulate | 0.007 | 0.097 | 0.036 | 0.027 | 0.167 | 15.564 | 1.557 | 2312 | 1172 |
| R Isthmus cingulate | 0.003 | 0.108 | 0.036 | 0.038 | 0.178 | 15.603 | 1.690 | 2306 | 1174 |
| **L Lateral occipital** | **<0.001** | **-0.125** | **0.036** | **-0.196** | **-0.054** | **-68.766** | **-1.452** | **2239** | **1152** |
| R Lateral occipital | 0.007 | -0.099 | 0.036 | -0.17 | -0.028 | -55.979 | -1.213 | 2257 | 1156 |
| L Lateral orbitofrontal | 0.427 | -0.029 | 0.036 | -0.099 | 0.042 | -7.421 | -0.293 | 2315 | 1169 |
| R Lateral orbitofrontal | 0.103 | 0.059 | 0.036 | -0.011 | 0.129 | 16.594 | 0.671 | 2312 | 1171 |
| L Lingual | 0.039 | -0.075 | 0.036 | -0.146 | -0.004 | -30.381 | -1.019 | 2251 | 1156 |
| R Lingual | 0.001 | -0.124 | 0.037 | -0.196 | -0.052 | -48.748 | -1.611 | 2214 | 1119 |
| L Medial orbitofrontal | 0.651 | -0.016 | 0.036 | -0.087 | 0.054 | -3.46 | -0.190 | 2295 | 1161 |
| R Medial orbitofrontal | 0.219 | -0.045 | 0.036 | -0.115 | 0.026 | -8.251 | -0.458 | 2291 | 1161 |
| L Middle temporal | 0.011 | -0.096 | 0.037 | -0.169 | -0.023 | -34.273 | -1.147 | 2109 | 1088 |
| R Middle temporal | 0.022 | -0.084 | 0.036 | -0.155 | -0.012 | -30.32 | -0.921 | 2234 | 1137 |
| L Paracentral | 0.003 | 0.108 | 0.036 | 0.037 | 0.178 | 19.065 | 1.435 | 2301 | 1170 |
| R Paracentral | 0.005 | 0.102 | 0.036 | 0.032 | 0.173 | 21.018 | 1.395 | 2291 | 1173 |
| L Parahippocampal | 0.315 | 0.036 | 0.036 | -0.034 | 0.107 | 3.699 | 0.547 | 2317 | 1171 |
| R Parahippocampal | 0.025 | 0.081 | 0.036 | 0.011 | 0.152 | 7.498 | 1.149 | 2303 | 1174 |
| L Pars opercularis | 0.706 | -0.014 | 0.036 | -0.084 | 0.057 | -3.265 | -0.206 | 2313 | 1173 |
| R Pars opercularis | 0.912 | -0.004 | 0.036 | -0.074 | 0.066 | -0.821 | -0.061 | 2308 | 1169 |
| L Pars orbitalis | 0.012 | -0.091 | 0.036 | -0.161 | -0.02 | -6.957 | -1.120 | 2305 | 1172 |
| R Pars orbitalis | 0.828 | 0.008 | 0.036 | -0.062 | 0.078 | 0.755 | 0.100 | 2309 | 1173 |
| L Pars triangularis | 0.786 | -0.01 | 0.036 | -0.08 | 0.061 | -1.792 | -0.145 | 2302 | 1174 |
| R Pars triangularis | 0.761 | 0.011 | 0.036 | -0.059 | 0.081 | 2.499 | 0.176 | 2295 | 1171 |
| L Pericalcarine | 0.04 | -0.076 | 0.037 | -0.149 | -0.004 | -18.444 | -1.357 | 2157 | 1114 |
| R Pericalcarine | 0.084 | -0.066 | 0.038 | -0.14 | 0.008 | -16.233 | -1.097 | 2086 | 1045 |
| L Postcentral | 0.298 | 0.038 | 0.037 | -0.033 | 0.11 | 15.779 | 0.385 | 2186 | 1133 |
| R Postcentral | 0.924 | -0.004 | 0.037 | -0.075 | 0.068 | -1.437 | -0.036 | 2182 | 1137 |
| L Posterior cingulate | 0.105 | 0.059 | 0.036 | -0.012 | 0.129 | 11.248 | 0.995 | 2319 | 1170 |
| R Posterior cingulate | 0.081 | 0.063 | 0.036 | -0.007 | 0.133 | 10.59 | 0.917 | 2316 | 1173 |
| L Precentral | 0.285 | 0.039 | 0.037 | -0.032 | 0.111 | 17.976 | 0.374 | 2214 | 1127 |
| R Precentral | 0.03 | 0.08 | 0.036 | 0.008 | 0.151 | 36.991 | 0.767 | 2231 | 1135 |
| L Precuneus | 0.305 | -0.037 | 0.036 | -0.107 | 0.033 | -14.996 | -0.407 | 2314 | 1172 |
| R Precuneus | 0.678 | 0.015 | 0.036 | -0.055 | 0.085 | 6.41 | 0.167 | 2314 | 1173 |
| L Rostral anterior cingulate | 0.628 | 0.018 | 0.036 | -0.053 | 0.089 | 2.677 | 0.327 | 2289 | 1142 |
| R Rostral anterior cingulate | 0.968 | -0.001 | 0.036 | -0.072 | 0.069 | -0.189 | -0.029 | 2294 | 1158 |
| L Rostral middle frontal | 0.232 | -0.043 | 0.036 | -0.114 | 0.027 | -27.117 | -0.498 | 2296 | 1172 |
| R Rostral middle frontal | 0.099 | -0.06 | 0.036 | -0.13 | 0.011 | -39.205 | -0.694 | 2312 | 1169 |
| L Superior frontal | 0.413 | -0.03 | 0.036 | -0.101 | 0.041 | -19.824 | -0.284 | 2268 | 1150 |
| R Superior frontal | 0.729 | 0.013 | 0.036 | -0.058 | 0.083 | 8.42 | 0.125 | 2271 | 1162 |
| L Superior parietal | 0.031 | -0.079 | 0.036 | -0.15 | -0.008 | -45.35 | -0.857 | 2243 | 1151 |
| R Superior parietal | 0.656 | -0.016 | 0.036 | -0.087 | 0.055 | -9.127 | -0.173 | 2262 | 1160 |
| L Superior temporal | 0.607 | -0.02 | 0.038 | -0.094 | 0.055 | -7.372 | -0.200 | 2027 | 1063 |
| R Superior temporal | 0.852 | 0.007 | 0.037 | -0.065 | 0.079 | 2.29 | 0.065 | 2158 | 1114 |
| L Supramarginal | 0.891 | 0.005 | 0.037 | -0.067 | 0.077 | 2.602 | 0.069 | 2191 | 1117 |
| R Supramarginal | 0.718 | 0.013 | 0.036 | -0.058 | 0.084 | 6.096 | 0.170 | 2225 | 1150 |
| L Temporal pole | 0.149 | -0.053 | 0.036 | -0.124 | 0.018 | -3.196 | -0.677 | 2227 | 1151 |
| R Temporal pole | 0.328 | -0.036 | 0.036 | -0.107 | 0.035 | -2.27 | -0.533 | 2229 | 1142 |
| L Transverse temporal | 0.009 | 0.094 | 0.036 | 0.023 | 0.164 | 6.944 | 1.554 | 2328 | 1175 |
| R Transverse temporal | 0.243 | 0.042 | 0.036 | -0.028 | 0.112 | 2.23 | 0.670 | 2328 | 1177 |

### Subcortical regions

The subcortical analysis revealed a significantly larger left thalamus (*d* = 0·17, 1·82%), smaller putamen bilaterally (left: *d* = −0·16, −2·01%; right: *d* = −0·16, −2·00%), and a smaller left amygdala (*d* = −0·15, −2·26%) in PD patients compared to controls (**Figure 4** and **Table 3**).

**Figure 4.**
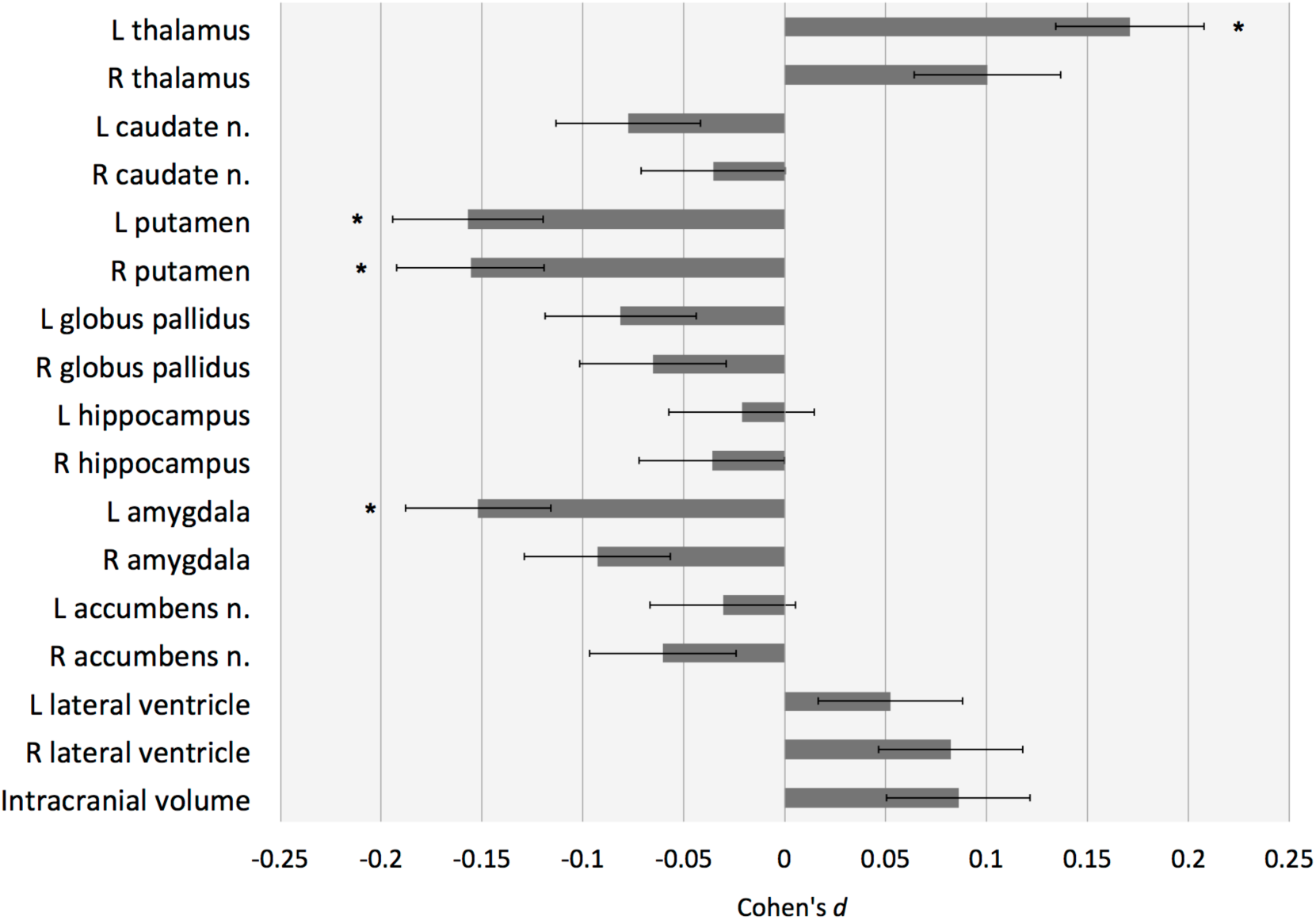
Subcortical volume findings for PD vs controls. Cohen’s *d* effect size estimates with error bars for differences in volume. Structures with *P*-values < 5·88 × 10^−5^ (i.e. 0·001/17 ROIs) are depicted as *. Abbreviations: L, left; R, right; ROI, region of interest; n., nucleus.

**Table 3:**
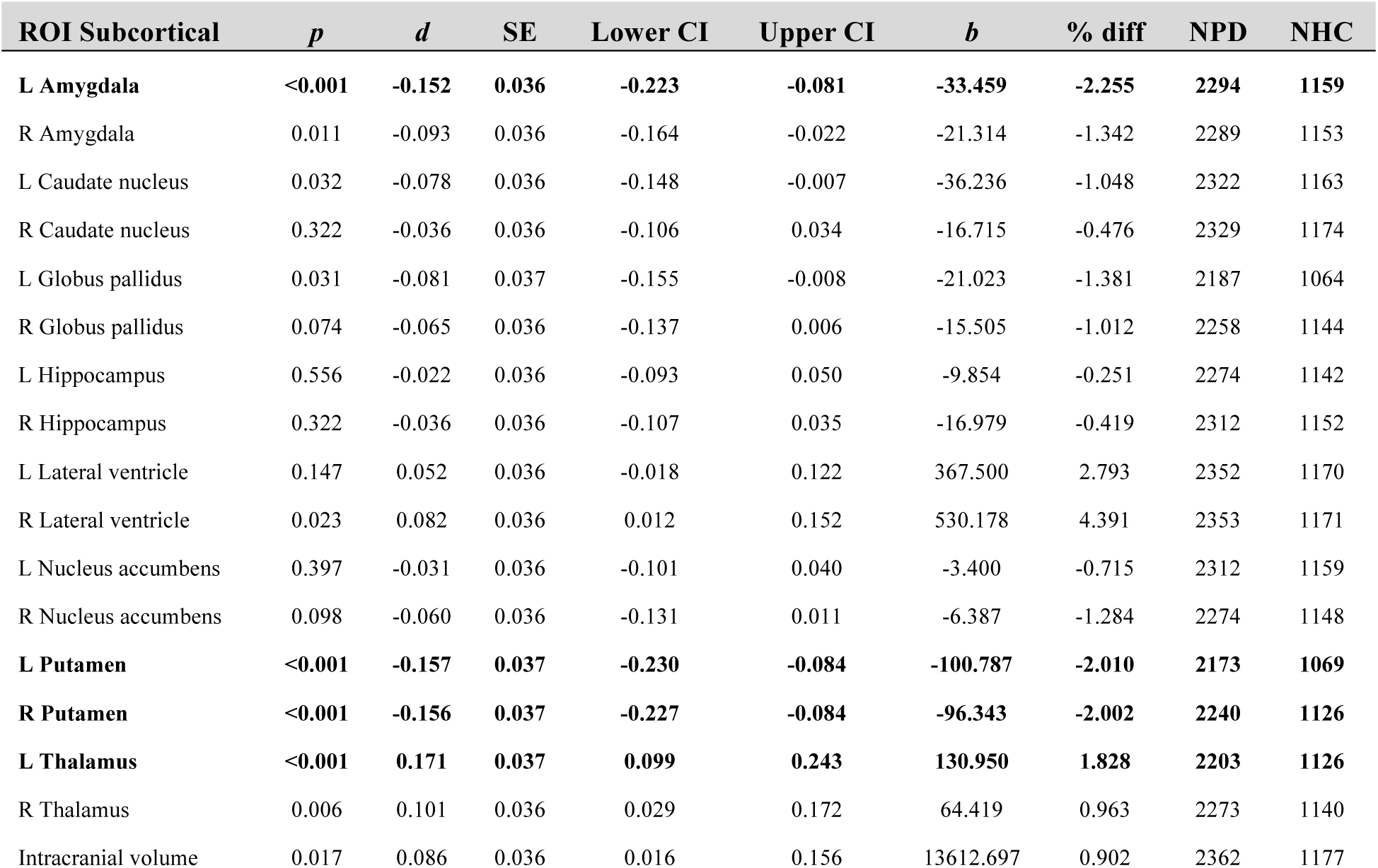
Subcortical volume in PD vs controls.

### Disease severity: Hoehn & Yahr stages

After the matching procedure, 437 stage 1 patients (847 controls), 941 stage 2 patients (908 controls), 263 stage 3 patients (502 controls), and 85 stage 4 & 5 patients (329 controls) were available for the analyses (**Tables S3a-d**). Included controls partially overlapped across stages. Wilcoxon signed-ranks tests revealed significant differences in illness duration and MoCA scores between all HY stage patient groups (**Figure S1**).

#### Cortical thickness

A summary of thickness results is shown in **Figure 5a**, **Table 4**, and **Table S6a-d**. Supplementary videos have been included to show thickness and surface area results across HY stages. Compared with controls, HY stage 1 patients showed a thinner left fusiform (*d* = −0·21, −1·31%) and inferior temporal cortex (*d* = −0·23, −1·43%) and right lateral occipital (*d* = −0·21, −1·56%), precuneus (*d* = −0·22, −1·47%), and inferior (*d* = −0·29, −1·92%) and superior parietal cortex (*d* = −0·25, −1·73%). HY stage 2 patients showed a thinner cortex in nine regions in the left hemisphere (*dmax* = −0·22, −2·55%; *dmin* = −0·17, −1·21%) and seven regions in the right hemisphere (*dmax* = −0·24, −2·00%; *dmin* = −0·17, −1·33%). HY stage 3 patients showed a thinner cortex in 14 regions in the left hemisphere (*dmax* = −0·49, −3·72%; *dmin* = −0·28, −2·26%) and 16 regions in the right hemisphere (*dmax* = −0·47, −4·78%; *dmin* = −0·28, −1·91%). HY stage 4 and 5 patients showed a thinner cortex in 13 regions in the left hemisphere (*dmax* = −0·76, −5·12%; *dmin* = −0·45, −2·79%) and 15 regions in the right hemisphere (*dmax* = −0·67, −5·68%; *dmin* = −0·44, −3·09%). Overall, the number of significantly different ROIs and their effect sizes were higher with increasing HY stage.

**Table 4:**
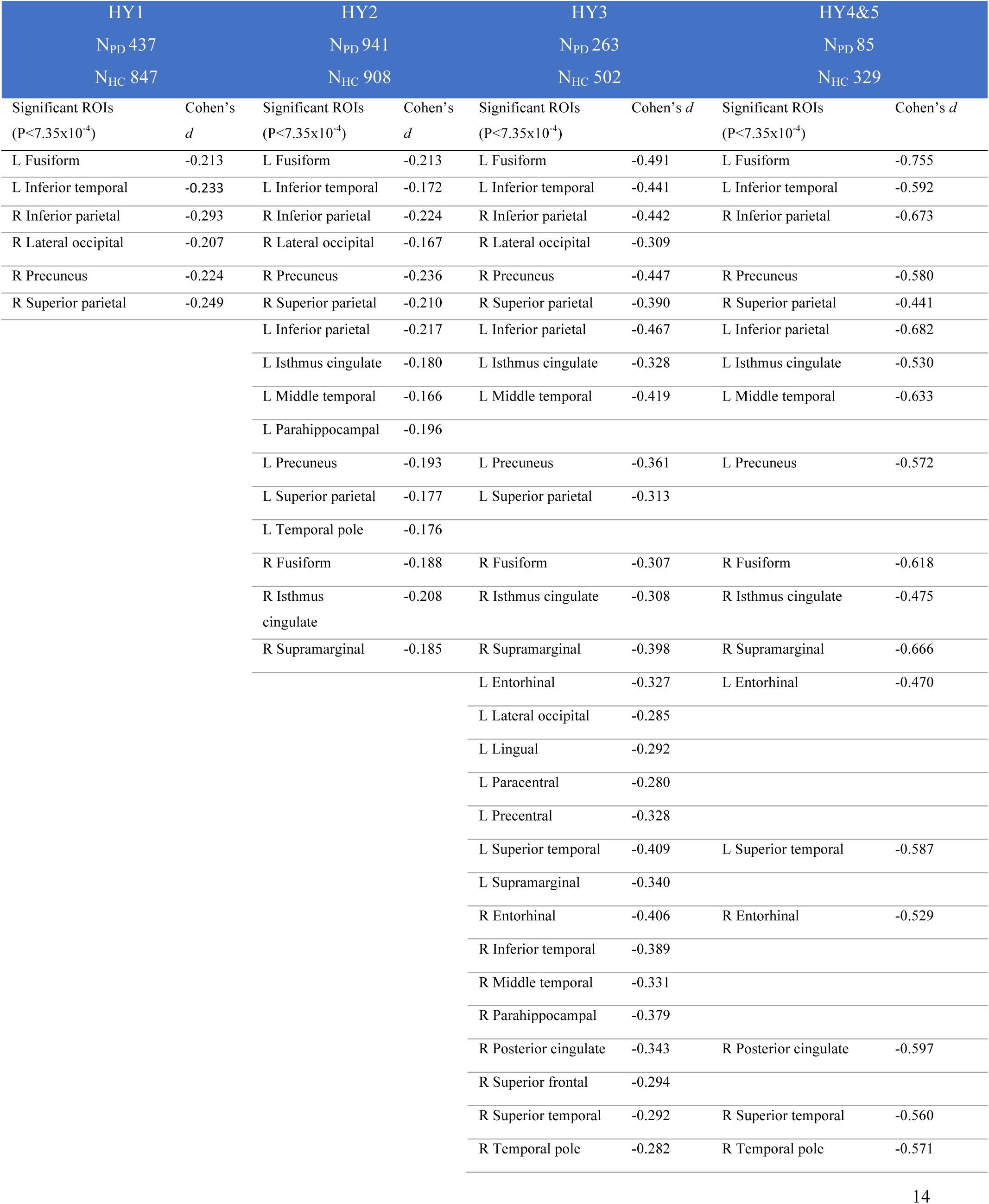
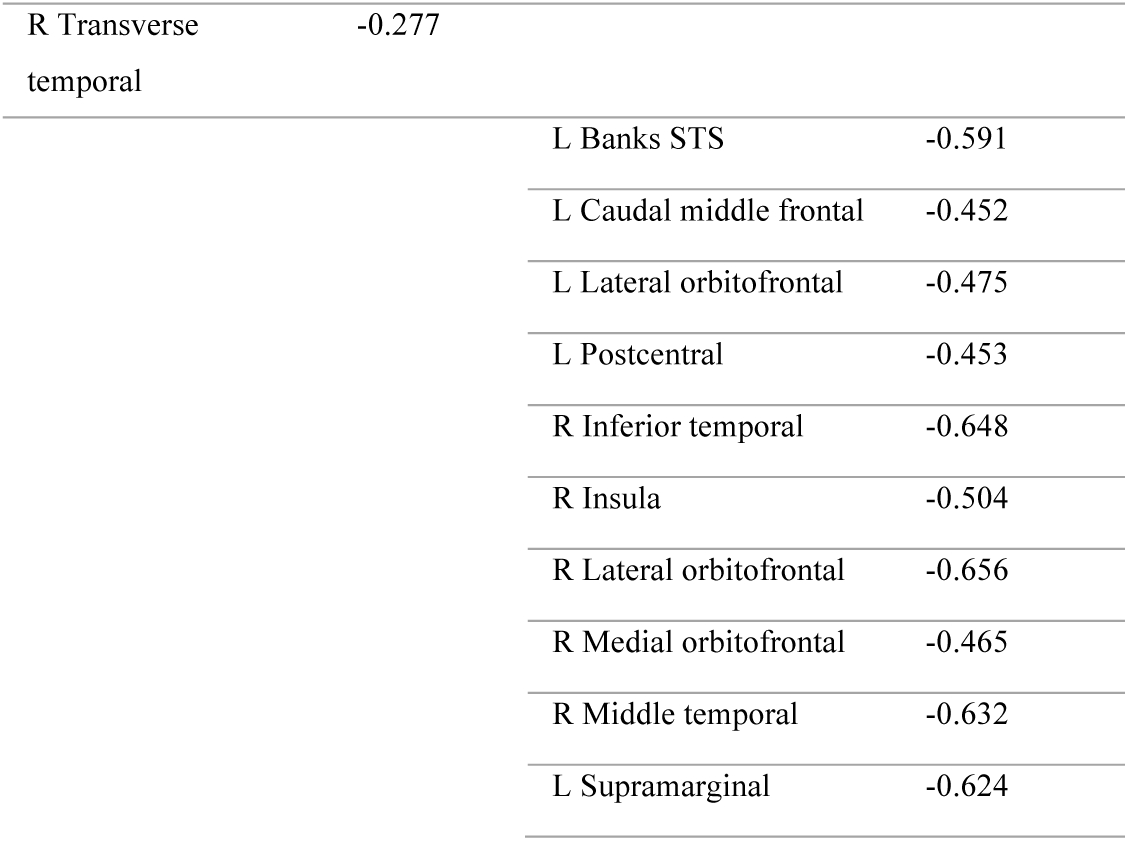
Summary of cortical thickness results across HY stages.

#### Cortical surface area

Compared to controls, HY stage 1 patients showed a smaller surface area of the left frontal pole (*d* = −0·25, −3·90%; **Figure 5b**, **Table 5**, and **Table S6e-h**). HY stage 2 patients showed a smaller surface area of the lingual cortex (left: *d* = −0·16, −2·30%; right: *d* = −0·19, −2·57%) and pericalcarine cortex (*d* = −0·18, −3·07%). HY stage 3 patients showed a smaller surface area in 9 regions in the left hemisphere (*dmax* = −0·35, −5·76%; *dmin* = −0·27, −3·34%) and 7 regions in the right hemisphere (*dmax* = −0·40, −4·69%; *dmin* = −0·27, −3·11%). HY stage 4 and 5 patients combined showed a smaller surface area of the precuneus (left: *d* = −0·49, −5·09%; right: *d* = −0·47, −4·89%), left inferior temporal (*d* = −0·47, −5·74%) and lateral occipital cortex (*d* = −0·48, −5·50%), and right middle temporal cortex (*d* = −0·45, −4·75%).

**Figure 5.**
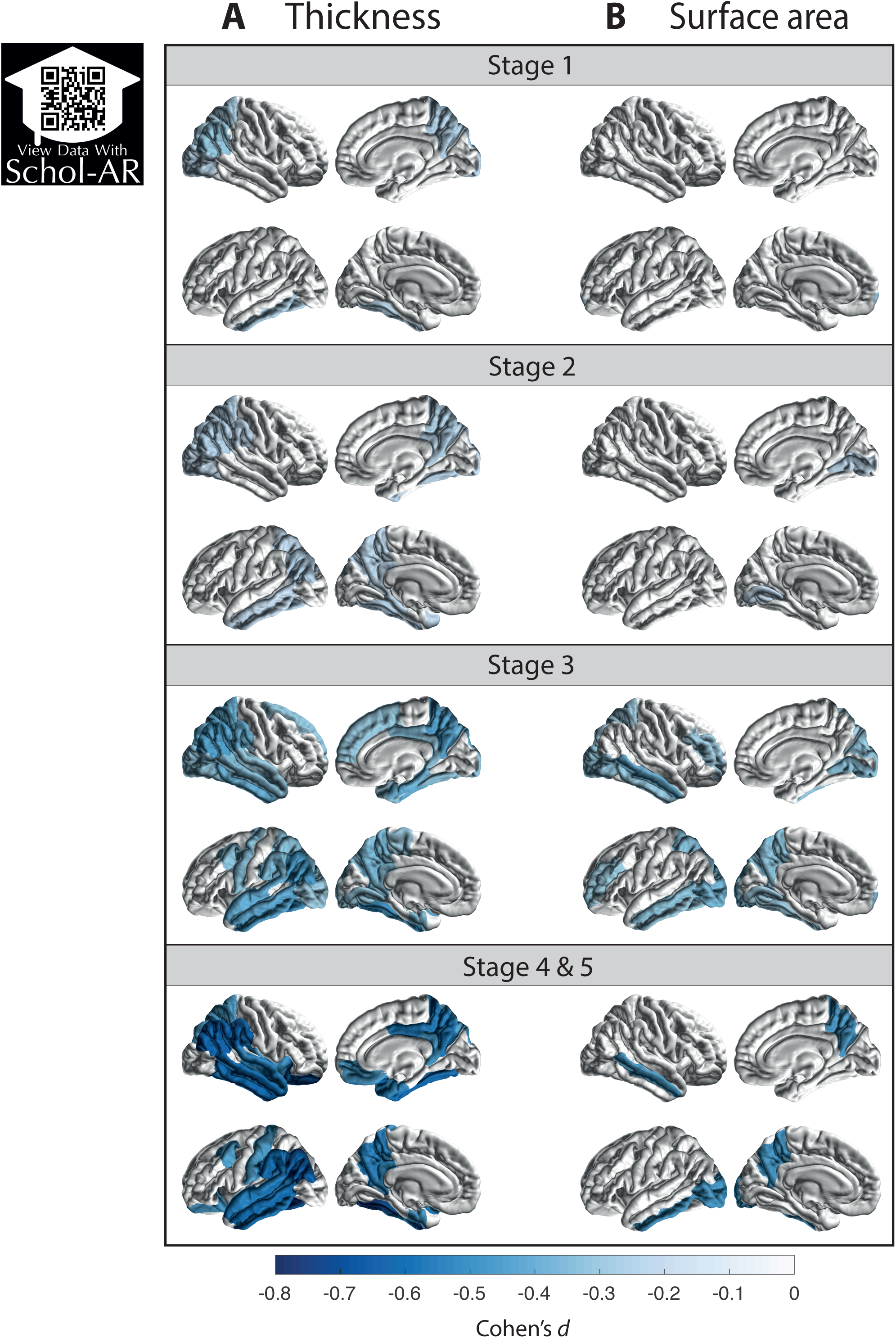
Cortical thickness and surface area group differences for PD groups, at different Hoehn & Yahr stages, versus age- and sex-matched controls. Cohen’s *d* effect size estimates for differences in (A) cortical thickness and (B) surface area. Cortical regions with P-values < 7·35 × 10-4 (i.e., 0·05/68 ROIs) are depicted in the heatmap colors. Abbreviation: ROI, region of interest.

**Table 5:** Summary of cortical surface area results across HY stages.

| HY1 |  | HY2 |  | HY3 |  | HY4&5 |  |
| --- | --- | --- | --- | --- | --- | --- | --- |
| N<sub>PD</sub> 437 |  | N<sub>PD</sub> 941 |  | N<sub>PD</sub> 263 |  | N<sub>PD</sub> 85 |  |
| N<sub>HC</sub> 847 |  | N<sub>HC</sub> 908 |  | N<sub>HC</sub> 502 |  | N<sub>HC</sub> 329 |  |
| Significant ROIs<br>($P < 7.35 \times 10^{-4}$) | Cohen's *d* | Significant ROIs<br>($P < 7.35 \times 10^{-4}$) | Cohen's *d* | Significant ROIs<br>($P < 7.35 \times 10^{-4}$) | Cohen's *d* | Significant ROIs<br>($P < 7.35 \times 10^{-4}$) | Cohen's *d* |
| L Frontal Pole | -0.250 |  |  | L Frontal Pole | -0.346 |  |  |
|  |  | L Lingual | -0.164 | L Lingual | -0.273 |  |  |
|  |  | R Lingual | -0.190 | R Lingual | -0.324 |  |  |
|  |  | R Pericalcarine | -0.184 | L Fusiform | -0.281 |  |  |
|  |  |  |  | L Inferior temporal | -0.280 | L Inferior temporal | -0.472 |
|  |  |  |  | L Lateral occipital | -0.351 | L Lateral occipital | -0.484 |
|  |  |  |  | L Middle temporal | -0.323 |  |  |
|  |  |  |  | L Precuneus | -0.318 | L Precuneus | -0.495 |
|  |  |  |  | L Rostral middle frontal | -0.277 |  |  |
|  |  |  |  | L Superior parietal | -0.328 |  |  |
|  |  |  |  | R Cuneus | -0.301 |  |  |
|  |  |  |  | R Inferior temporal | -0.288 |  |  |
|  |  |  |  | R Lateral occipital | -0.310 |  |  |
|  |  |  |  | R Middle temporal | -0.395 | R Middle temporal | -0.454 |
|  |  |  |  | R Rostral middle frontal | -0.269 |  |  |
|  |  |  |  | R Superior parietal | -0.273 |  |  |
|  |  |  |  |  |  | R Precuneus | -0.470 |

#### Subcortical volumes

Results of the subcortical analysis are depicted in **Figure 6**, **Table 6**, and **Table S7a-d**. Compared to controls, HY stage 1 patients showed a significantly larger left thalamus (*d* = 0·19, 2·14%). HY stage 2 patients showed smaller bilateral amygdalae (left: *d* = −0·18, −2·98%; right:*d* = −0·18, −2·29%) and smaller putamen (left: *d* = −0·19, −2·44%; right: *d* = −0·15, −2·47%). At HY stage 3, patients showed smaller amygdalae (left: *d* = −0·51, −8·69%; right: *d* = −0·37, −6·35%), putamen (left: *d* = −0·33, −4·67%; right: *d* = −0·30, −4·14%), hippocampi (left: *d* = −0·27, −3·47%; right: *d* = −0·32, −4·33%), and left caudate nucleus (*d* = −0·25, −3·66%). Finally,HY stage 4 and 5 patients showed smaller amygdalae (left: *d* = −0·78, −11·60%; right: *d* = −0·69, −10·06%), hippocampi (left: *d* = −0·60, −6·72%; right: *d* = −0·71, −8·02%), putamen (left: *d* = −0·52, −6·27%; right: *d* = −0·57, −7·19%), left caudate nucleus (*d* = −0·39, −4·94%) and *globus pallidus* (*d* = −0·40, −6·67%), and right accumbens (*d* = −0·62, −13·04%). The lateral ventricles were larger in PD (left: *d* = 0·41, −18·66%; right: *d* = 0·59, −27·87%).

**Table 6:** Summary of subcortical volume results across HY stages.

| HY1 |  | HY2 |  | HY3 |  | HY4&5 |  |
| --- | --- | --- | --- | --- | --- | --- | --- |
| N<sub>PD</sub> 437 |  | N<sub>PD</sub> 941 |  | N<sub>PD</sub> 263 |  | N<sub>PD</sub> 85 |  |
| N<sub>HC</sub> 847 |  | N<sub>HC</sub> 908 |  | N<sub>HC</sub> 502 |  | N<sub>HC</sub> 329 |  |
| Significant ROIs<br>($P < 6.25 \times 10^{-5}$) | Cohen's<br>$d$ | Significant<br>ROIs<br>($P < 6.25 \times 10^{-5}$) | Cohen's<br>$d$ | Significant ROIs<br>($P < 6.25 \times 10^{-5}$) | Cohen's<br>$d$ | Significant ROIs<br>($P < 6.25 \times 10^{-5}$) | Cohen's<br>$d$ |
| L Thalamus | 0.191 |  |  |  |  |  |  |
|  |  | L Amygdala | -0.191 | L Amygdala | -0.513 | L Amygdala | -0.778 |
|  |  | L Putamen | -0.179 | L Putamen | -0.335 | L Putamen | -0.522 |
|  |  | R Amygdala | -0.149 | R Amygdala | -0.374 | R Amygdala | -0.694 |
|  |  | R Putamen | -0.183 | R Putamen | -0.297 | R Putamen | -0.568 |
|  |  |  |  | L Caudate<br>nucleus | -0.252 | L Caudate nucleus | -0.388 |
|  |  |  |  | L Hippocampus | -0.268 | L Hippocampus | -0.605 |
|  |  |  |  | R Hippocampus | -0.324 | R Hippocampus | -0.706 |
|  |  |  |  |  |  | L Globus pallidus | -0.398 |
|  |  |  |  |  |  | L Lateral ventricle | 0.407 |
|  |  |  |  |  |  | R Lateral ventricle | 0.588 |
|  |  |  |  |  |  | R Nucleus accumbens | -0.619 |

**Figure 6.**
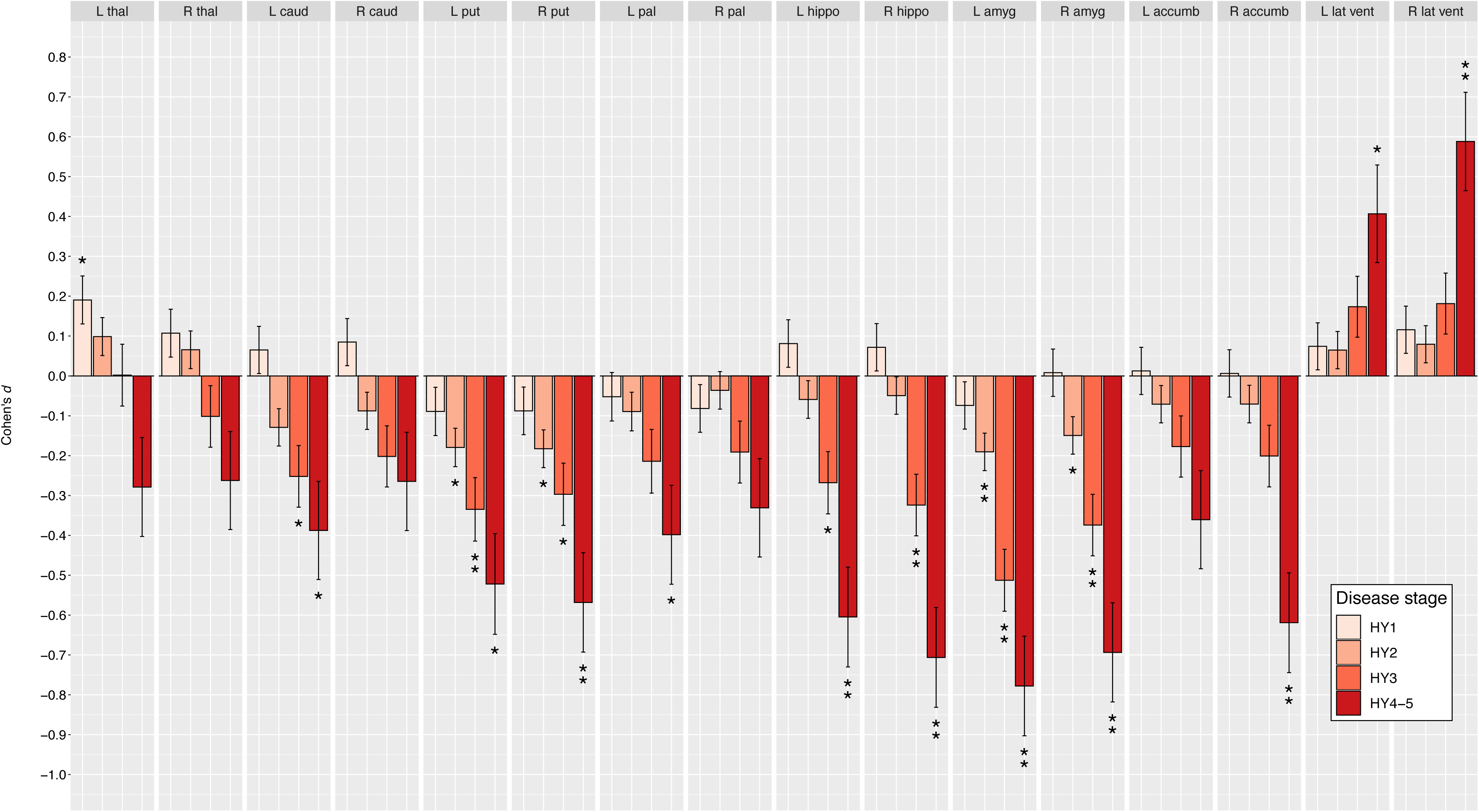
Subcortical group differences for PD groups, at different Hoehn & Yahr stages, versus age- and sex- matched controls. Cohen’s d effect size estimates are shown for differences in mean volume. Cortical regions with P-values < 3·13 × 10^-3^ (i.e. 0·05/16 ROIs) are depicted as * and structures with P-values < 6·25×10^-5^ as ** (i.e. 0·001/16 ROIs). Abbreviation: ROI, region of interest; L, left; R, right; thal, thalamus; amyg, amygdala; caud, caudate nucleus; hippo, hippocampus; accumb, accumbens nucleus; put, putamen; pal, globus pallidus; lat vent, lateral ventricle.

#### Medication status

For the analyses, 133 medicated and 196 non-medicated HY stage 1 patients, and 573 medicated and 229 non-medicated HY stage 2 patients were available (**Tables S4a,b**). Analyses on HY stage 3 and HY stage 4 and 5 were not performed, due to limited availability of non-medicated patients (eight and four, respectively; **Table S3c,d**). All results are shown in the supplementary materials (**Tables S8a-f**). The medication analysis revealed no significant subcortical volume differences between medicated and unmedicated patients in HY stage 1. The HY stage 2 analysis showed significantly larger lateral ventricles in medicated patients (left: *d* = 0·28; right: *d* = 0·25). We found no significant differences between medicated and unmedicated PD patients within HY stage 1 and 2 for any cortical measures.

#### Sex by diagnosis interaction

Results are shown in the supplementary material (**Tables S10a-o**). We found no significant interactions between sex and diagnosis in the whole sample, and at any HY stage, for cortical thickness, cortical surface area, and subcortical volume.

### MoCA

#### Cortex

A total of 1,044 patients had MoCA scores available for analysis (**Table S5**). Thickness results are depicted in **Figure 7a** and **Table 7a**. The analysis revealed a significant positive association between MoCA scores and cortical thickness in 16 regions in the left hemisphere (*t*max = 5·84, *t*min = 3·50) and 12 regions in the right hemisphere (*t*max = 5·42, *t*min = 3·46). Surface area results are shown in **Figure 6b** and **Table 7b**. We found a significant positive association between MoCA scores and cortical surface area in the left *pars opercularis* (*t* = 4·21), precuneus (*t* = 3·55), and superior parietal cortex (*t* = 3·55) and the right inferior parietal (*t* = 4·52) and superior parietal cortex (*t* = 3·52). MoCA PD patients versus controls results are depicted in **Tables S9a,b**.

**Figure 7.**
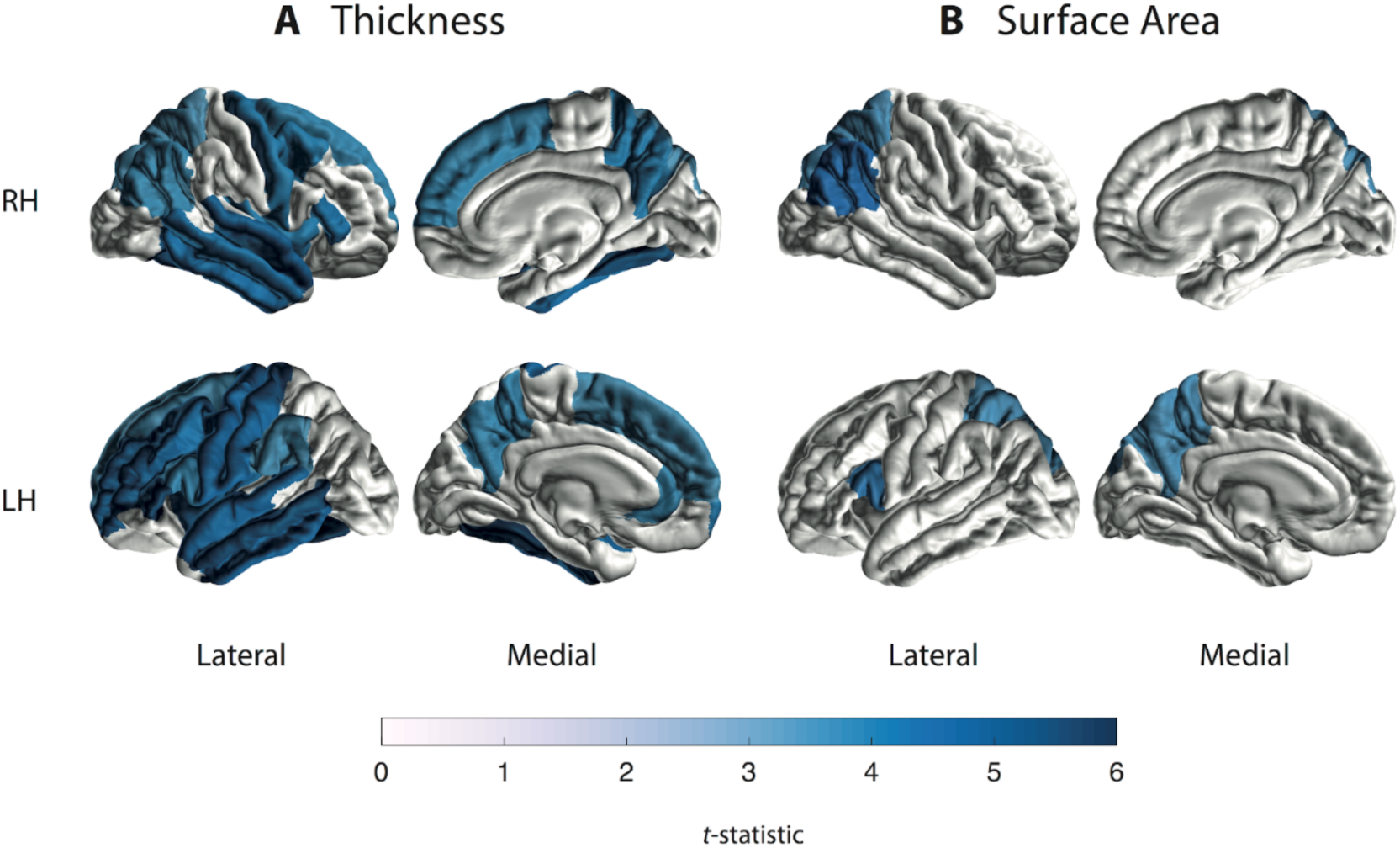
Cortical thickness and surface area findings for the MoCA regression. *t*-statistic estimates for the associations with (A) cortical thickness and (B) cortical surface area. Cortical regions with *P*-values < 7·35 × 10^−4^ (i.e., 0·05/68 ROIs) are depicted in the heatmap colors. Higher MoCA scores denote better cognitive performance. Abbreviations: RH, right hemisphere; LH, left hemisphere.

**Table 7a:** MoCA score and cortical thickness regression.

| ROI Thickness | *p* | *t* | **b** | NPD |
| --- | --- | --- | --- | --- |
| L Banks STS | 0.001 | 3.323 | 0.006 | 947 |
| R Banks STS | 0.006 | 2.771 | 0.005 | 1013 |
| L Caudal anterior cingulate | 0.281 | 1.079 | 0.003 | 1034 |
| R Caudal anterior cingulate | 0.585 | 0.547 | 0.001 | 1033 |
| **L Caudal middle frontal** | **<0.001** | **4.361** | **0.007** | **1042** |
| **R Caudal middle frontal** | **<0.001** | **4.109** | **0.006** | **1042** |
| L Cuneus | 0.057 | 1.906 | 0.003 | 946 |
| R Cuneus | 0.247 | 1.159 | 0.002 | 937 |
| L Entorhinal | 0.081 | 1.749 | 0.006 | 1025 |
| R Entorhinal | 0.056 | 1.913 | 0.006 | 1019 |
| **L Frontal pole** | **<0.001** | **4.127** | **0.011** | **1039** |
| R Frontal pole | 0.002 | 3.040 | 0.008 | 1038 |
| **L Fusiform** | **<0.001** | **5.836** | **0.009** | **1020** |
| **R Fusiform** | **<0.001** | **5.415** | **0.009** | **1027** |
| L Inferior parietal | 0.001 | 3.323 | 0.005 | 1014 |
| **R Inferior parietal** | **<0.001** | **3.676** | **0.005** | **1012** |
| **L Inferior temporal** | **<0.001** | **4.698** | **0.007** | **999** |
| **R Inferior temporal** | **<0.001** | **4.382** | **0.007** | **1015** |
| **L Insula** | **<0.001** | **4.220** | **0.007** | **1039** |
| **R Insula** | **<0.001** | **4.070** | **0.008** | **1036** |
| L Isthmus cingulate | 0.058 | 1.894 | 0.004 | 1039 |
| R Isthmus cingulate | 0.003 | 3.030 | 0.006 | 1039 |
| L Lateral occipital | 0.281 | 1.078 | 0.001 | 1013 |
| R Lateral occipital | 0.010 | 2.574 | 0.004 | 1015 |
| L Lateral orbitofrontal | 0.009 | 2.629 | 0.004 | 1033 |
| R Lateral orbitofrontal | 0.002 | 3.138 | 0.005 | 1034 |
| L Lingual | 0.002 | 3.161 | 0.004 | 1006 |
| R Lingual | 0.008 | 2.660 | 0.003 | 989 |
| L Medial orbitofrontal | 0.004 | 2.891 | 0.005 | 1027 |
| R Medial orbitofrontal | 0.009 | 2.633 | 0.005 | 1030 |
| **L Middle temporal** | **<0.001** | **5.489** | **0.010** | **949** |
| **R Middle temporal** | **<0.001** | **4.306** | **0.007** | **1013** |
| L Paracentral | 0.003 | 2.996 | 0.005 | 1036 |
| R Paracentral | 0.002 | 3.106 | 0.005 | 1035 |
| L Parahippocampal | 0.103 | 1.634 | 0.005 | 1038 |
| R Parahippocampal | 0.002 | 3.172 | 0.008 | 1038 |
| **L Pars opercularis** | **<0.001** | **3.802** | **0.006** | **1033** |
| R Pars opercularis | 0.001 | 3.252 | 0.005 | 1031 |
| L Pars orbitalis | 0.064 | 1.852 | 0.004 | 1033 |
| R Pars orbitalis | 0.029 | 2.191 | 0.004 | 1032 |
| **L Pars triangularis** | **<0.001** | **5.532** | **0.008** | **1031** |
| **R Pars triangularis** | **<0.001** | **4.267** | **0.006** | **1030** |
| L Pericalcarine | 0.051 | 1.951 | 0.003 | 947 |
| R Pericalcarine | 0.103 | 1.633 | 0.002 | 920 |
| **L Postcentral** | **<0.001** | **4.561** | **0.006** | **1003** |
| R Postcentral | 0.001 | 3.239 | 0.004 | 1004 |
| L Posterior cingulate | 0.002 | 3.052 | 0.005 | 1040 |
| R Posterior cingulate | 0.003 | 2.950 | 0.005 | 1039 |
| **L Precentral** | **<0.001** | **4.602** | **0.008** | **1026** |
| **R Precentral** | **<0.001** | **4.099** | **0.007** | **1029** |
| **L Precuneus** | **<0.001** | **3.764** | **0.005** | **1041** |
| **R Precuneus** | **<0.001** | **4.118** | **0.006** | **1040** |
| **L Rostral anterior cingulate** | **<0.001** | **3.773** | **0.009** | **1029** |
| R Rostral anterior cingulate | 0.533 | 0.624 | 0.002 | 1033 |
| **L Rostral middle frontal** | **<0.001** | **5.076** | **0.006** | **1034** |
| R Rostral middle frontal | 0.002 | 3.173 | 0.004 | 1034 |
| **L Superior frontal** | **<0.001** | **3.548** | **0.005** | **1019** |
| **R Superior frontal** | **<0.001** | **3.771** | **0.005** | **1020** |
| L Superior parietal | 0.002 | 3.033 | 0.004 | 1011 |
| **R Superior parietal** | **0.001** | **3.458** | **0.005** | **1025** |
| **L Superior temporal** | **<0.001** | **4.811** | **0.008** | **910** |
| **R Superior temporal** | **<0.001** | **4.891** | **0.008** | **987** |
| **L Supramarginal** | **<0.001** | **3.497** | **0.005** | **1005** |
| R Supramarginal | 0.001 | 3.366 | 0.005 | 1013 |
| L Temporal pole | 0.007 | 2.719 | 0.009 | 978 |
| R Temporal pole | 0.008 | 2.654 | 0.009 | 1007 |
| L Transverse temporal | 0.006 | 2.748 | 0.006 | 1044 |
| R Transverse temporal | 0.003 | 2.979 | 0.007 | 1043 |

**Table 7b:** MoCA score and cortical surface area regression.

| ROI Surface Area | *p* | *t* | *b* | NPD |
| --- | --- | --- | --- | --- |
| L Banks STS | 0.005 | 2.793 | 4.191 | 947 |
| R Banks STS | 0.047 | 1.991 | 2.607 | 1013 |
| L Caudal anterior cingulate | 0.020 | 2.324 | 3.081 | 1034 |
| R Caudal anterior cingulate | 0.071 | 1.807 | 2.709 | 1033 |
| L Caudal middle frontal | 0.038 | 2.075 | 6.660 | 1043 |
| R Caudal middle frontal | 0.247 | 1.159 | 3.638 | 1043 |
| L Cuneus | 0.847 | 0.192 | 0.368 | 946 |
| R Cuneus | 0.435 | 0.782 | 1.574 | 937 |
| L Entorhinal | 0.966 | -0.043 | -0.037 | 1025 |
| R Entorhinal | 0.803 | 0.249 | 0.196 | 1020 |
| L Frontal pole | 0.206 | 1.264 | 0.373 | 1039 |
| R Frontal pole | 0.039 | 2.063 | 0.808 | 1038 |
| L Fusiform | 0.379 | 0.880 | 2.855 | 1021 |
| R Fusiform | 0.237 | 1.183 | 3.703 | 1026 |
| L Inferior parietal | 0.009 | 2.623 | 13.054 | 1014 |
| **R Inferior parietal** | **<0.001** | **4.630** | **25.999** | **1012** |
| L Inferior temporal | 0.180 | 1.342 | 5.318 | 999 |
| R Inferior temporal | 0.078 | 1.764 | 6.122 | 1015 |
| L Insula | 0.859 | 0.177 | 0.349 | 1039 |
| R Insula | 0.680 | -0.413 | -0.966 | 1036 |
| L Isthmus cingulate | 0.070 | 1.813 | 2.565 | 1039 |
| R Isthmus cingulate | 0.825 | -0.222 | -0.297 | 1039 |
| L Lateral occipital | 0.713 | 0.368 | 1.846 | 1013 |
| R Lateral occipital | 0.021 | 2.318 | 12.369 | 1015 |
| L Lateral orbitofrontal | 0.143 | 1.467 | 3.450 | 1034 |
| R Lateral orbitofrontal | 0.149 | 1.444 | 3.719 | 1035 |
| L Lingual | 0.150 | 1.442 | 5.196 | 1006 |
| R Lingual | 0.224 | 1.216 | 4.366 | 989 |
| L Medial orbitofrontal | 0.049 | 1.970 | 3.785 | 1026 |
| R Medial orbitofrontal | 0.936 | 0.080 | 0.137 | 1029 |
| L Middle temporal | 0.041 | 2.047 | 7.010 | 949 |
| R Middle temporal | 0.019 | 2.349 | 7.813 | 1013 |
| L Paracentral | 0.793 | -0.262 | -0.408 | 1036 |
| R Paracentral | 0.559 | -0.585 | -1.161 | 1035 |
| L Parahippocampal | 0.227 | 1.209 | 1.131 | 1038 |
| R Parahippocampal | 0.014 | 2.454 | 2.180 | 1038 |
| **L Pars opercularis** | **<0.001** | **4.284** | **9.029** | **1033** |
| R Pars opercularis | 0.051 | 1.957 | 3.579 | 1031 |
| L Pars orbitalis | 0.002 | 3.081 | 2.209 | 1033 |
| R Pars orbitalis | 0.035 | 2.110 | 1.875 | 1032 |
| L Pars triangularis | 0.042 | 2.032 | 3.388 | 1031 |
| R Pars triangularis | 0.119 | 1.559 | 3.250 | 1030 |
| L Pericalcarine | 0.786 | -0.271 | -0.667 | 947 |
| R Pericalcarine | 0.815 | -0.234 | -0.550 | 920 |
| L Postcentral | 0.048 | 1.978 | 7.442 | 1003 |
| R Postcentral | 0.032 | 2.148 | 8.188 | 1004 |
| L Posterior cingulate | 0.096 | 1.666 | 3.214 | 1040 |
| R Posterior cingulate | 0.194 | 1.301 | 2.009 | 1039 |
| L Precentral | 0.181 | 1.337 | 5.567 | 1026 |
| R Precentral | 0.004 | 2.920 | 12.546 | 1029 |
| **L Precuneus** | **<0.001** | **3.679** | **13.593** | **1041** |
| R Precuneus | 0.050 | 1.964 | 7.794 | 1040 |
| L Rostral anterior cingulate | 0.891 | -0.137 | -0.209 | 1029 |
| R Rostral anterior cingulate | 0.451 | 0.754 | 0.919 | 1033 |
| L Rostral middle frontal | 0.190 | 1.311 | 7.742 | 1034 |
| R Rostral middle frontal | 0.164 | 1.394 | 8.401 | 1034 |
| L Superior frontal | 0.953 | 0.059 | 0.379 | 1020 |
| R Superior frontal | 0.113 | 1.588 | 10.358 | 1020 |
| **L Superior parietal** | **<0.001** | **3.667** | **19.248** | **1011** |
| **R Superior parietal** | **<0.001** | **3.638** | **18.567** | **1025** |
| L Superior temporal | 0.048 | 1.984 | 7.430 | 910 |
| R Superior temporal | 0.071 | 1.808 | 5.671 | 987 |
| **L Supramarginal** | **<0.001** | **3.346** | **16.837** | **1005** |
| R Supramarginal | 0.119 | 1.560 | 6.927 | 1013 |
| L Temporal pole | 0.368 | 0.900 | 0.548 | 978 |
| R Temporal pole | 0.312 | -1.012 | -0.598 | 1007 |
| L Transverse temporal | 0.020 | 2.331 | 1.656 | 1044 |
| R Transverse temporal | 0.026 | 2.237 | 1.052 | 1043 |

#### Subcortical regions

A total of 1,065 patients had MoCA scores available for analysis (**Table S5**). Volume results are depicted in **Figure 8** and **Table 8**. The analysis revealed a significant positive association between MoCA scores and the volume of the hippocampus (left: *t* = 4·11; right: *t* = 4·67) and the amygdala (left: *t* = 5·33; right: *t* = 4·14) bilaterally. In addition, we found a negative association between MoCA scores and lateral ventricular volume bilaterally (left: *t* = −4·93; right: *t* = −4·19). MoCA PD patients versus controls results are depicted in **Table S9c**.

**Figure 8.**
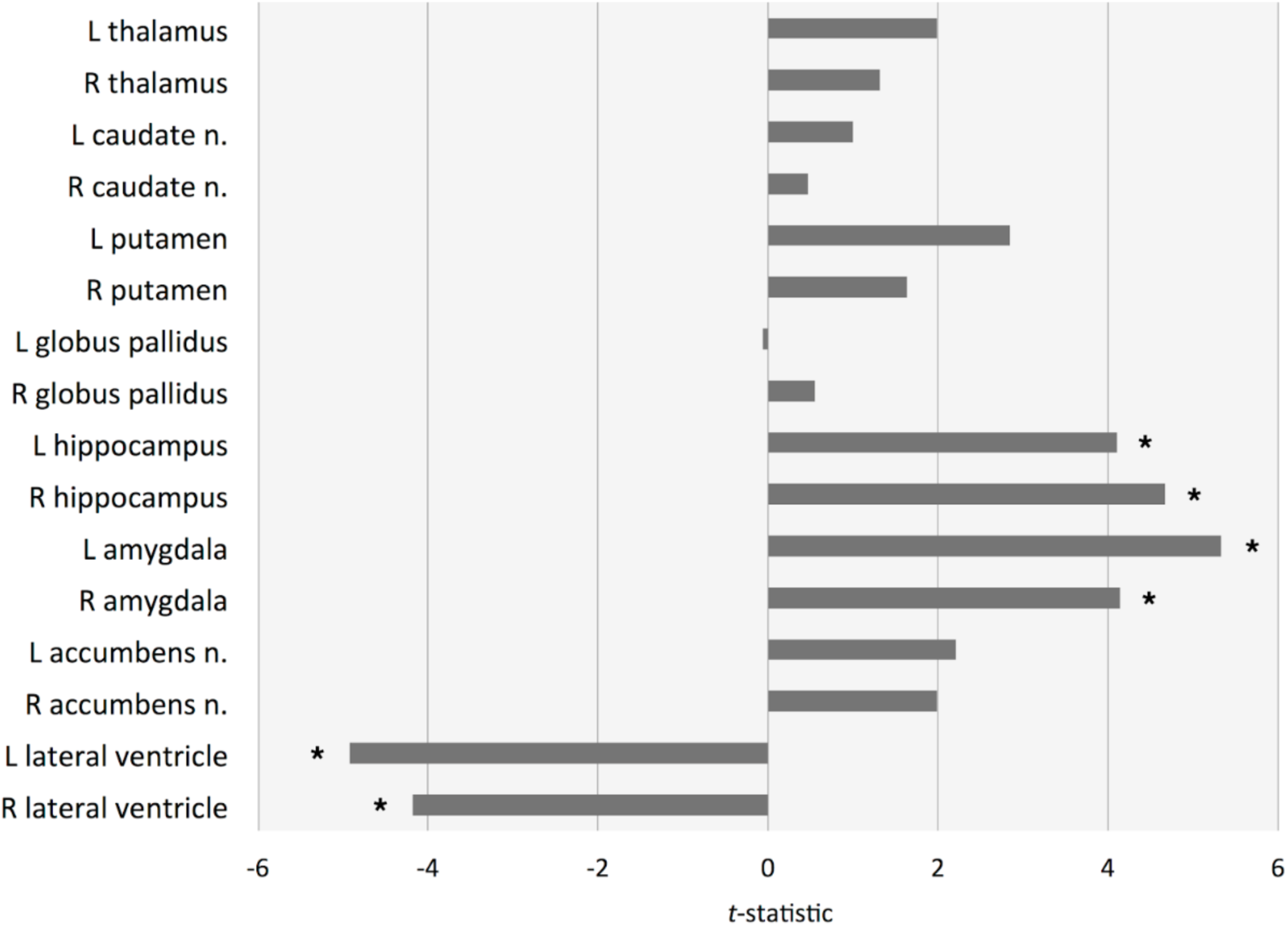
Subcortical volume findings for the MoCA regression. *t*-statistic estimates for the associations with ROI volume. Structures with *P*-values < 6·25 × 10^−5^ (i.e. 0·001/16 ROIs) are depicted as *. Higher MoCA scores denote better cognitive performance. Abbreviations: ROI, region of interest; n., nucleus.

**Table 8:** MoCA score and subcortical volume regression.

| **ROI Subcortical** | ***p*** | ***t*** | ***b*** | **NPD** |
| --- | --- | --- | --- | --- |
| **L Amygdala** | **<0.001** | **5.388** | **10.68** | **1059** |
| **R Amygdala** | **<0.001** | **4.224** | **8.75** | **1061** |
| L Caudate nucleus | 0.272 | 1.100 | 5.00 | 1056 |
| R Caudate nucleus | 0.556 | 0.589 | 2.73 | 1057 |
| L Globus pallidus | 0.988 | 0.014 | 0.03 | 1035 |
| R Globus pallidus | 0.527 | 0.632 | 1.37 | 1052 |
| **L Hippocampus** | **<0.001** | **4.181** | **16.76** | **1044** |
| **R Hippocampus** | **<0.001** | **4.743** | **19.94** | **1058** |
| **L Lateral ventricle** | **<0.001** | **-4.750** | **-337.85** | **1063** |
| **R Lateral ventricle** | **<0.001** | **-4.003** | **-262.67** | **1063** |
| L Nucleus accumbens | 0.025 | 2.239 | 2.43 | 1060 |
| R Nucleus accumbens | 0.044 | 2.012 | 2.05 | 1061 |
| L Putamen | 0.003 | 2.928 | 16.82 | 1036 |
| R Putamen | 0.090 | 1.699 | 9.52 | 1051 |
| L Thalamus | 0.034 | 2.120 | 14.75 | 1021 |
| R Thalamus | 0.143 | 1.465 | 8.62 | 1035 |

## Discussion

### Main findings

This is the largest collaborative MRI study to date to investigate PD-related alterations in cortical and subcortical brain morphology. We found lower cortical thickness, on average, in patients compared to controls across all HY disease stages, showing more pronounced alterations with disease severity. In the subcortex, a larger left thalamus in patients in stage 1 was followed by smaller putamen and amygdalae bilaterally in stage 2 and onwards. In addition, late-stage patients showed smaller hippocampi, left caudate nucleus, left *globus pallidus*, and right accumbens, while larger lateral ventricles were observed. Finally, we found that poorer cognitive cognitive performance was associated with widespread cortical atrophy as well as volume loss in core limbic structures.

### HY and disease staging

The HY staging scale has been developed to assess disease severity (i.e., degree of disability and impairment) of parkinsonism in the context of lateralization of motor symptoms and compromised balance/gait.^6^ The HY stages have been shown to be useful for tracking disease progression,^13^ although the relationship between motor and cognitive symptom development has not been fully elucidated. Generally, both domains tend to worsen during the disease course, with a dementia prevalence up to 80% in the final stages of PD, in addition to severe movement disabilities.^14^ Our cortical and subcortical findings are strongly in line with an ongoing neurodegenerative process; each HY increment largely replicates the previous stage with additional implicated regions, emphasized by longer illness duration and poorer cognitive performance in patients.

### Cognitive features

Thinner posterior and temporal cortical regions have been linked to cognitive impairment in the early symptomatic stages of PD.^15^ Indeed, we found that poorer cognition was associated with thinning in the parietal and inferior temporal regions, which may contingent upon the cortical alterations as demonstrated in HY stage 1 and 2 patients. We may, however, assume the vast majority of early-stage patients were cognitively normal,^16^ which would fit the notion that temporal and parietal degeneration may precede cognitive decline.^15^ In addition, the early implication of the occipital cortex may relate to a compromised visual system, in line with visual disturbances that are frequently reported in early-stage PD patients.^17^ The diffuse pattern of thinner cortex across the cortex alongside smaller hippocampi and amygdalae in HY stage 3 patients is consistent with the more advanced symptomatic stages associated with PD dementia;^18^ we found those regions accordingly linked to worse cognitive performance. Patients in the final stages proceeded showing the degenerative pattern with enlarged lateral ventricles - highlighting the severe atrophy in surrounding and adjacent structures, such as the hippocampus.

### Motor features

An estimated 80% of striatal dopaminergic neurons are lost at the time of motor symptom onset in PD.^19^ Dysfunction of the nigrostriatal pathway is associated with motor symptoms and is emphasized by reduced activity in the putamen, forming a well-established biomarker of early-stage PD.^20^ In line with this, we observed lower putamen volumes that are indicative of early abnormal atrophy in PD; the symmetry in HY stage 2 is appropriate to the transition from unilateral to bilateral motor impairment. Striatal degeneration is further highlighted by atrophy of the caudate nucleus in HY stage 3 and onwards. The *globus pallidus* appeared robust to volume loss until final stages; considering the role of pallidal dopamine depletion in the production of tremor in PD,^21^ this may be unexpected. It is noteworthy that the *globus pallidus* in particular tends to show poor contrast on T1-weighted scans,^22^ which may influence adequate segmentation. A counterintuitive finding in view of neurodegeneration was the higher volume of the left thalamus in HY stage 1 patients compared to controls. This did not seem to be explained by the use of dopaminergic medication. However, due to the complex structural and functional organisation of the different thalamic nuclei, a straightforward interpretation based on our global volume findings is challenging. Studies that incorporated atlas-based methods for segmentation of subnuclei or vertex-wise methods for surface deformation have reported local differences to suggest both atrophic and hypertrophic regions within the thalamus.^23,24^ It is proposed that hyperactivity in the cerebellothalamic circuit underlies Parkinson tremor,^21^ which may account for temporary thalamic hypertrophy. Follow-up research focused on regional morphology within the thalamus may shed light on localized alterations related to disease severity.

### Relation to Braak’s staging model

According to Braak’s staging model,^25^ Lewy body pathology spreads in an ascending fashion from brainstem regions towards the subcortex, finally reaching the neocortex through the mesocortex. It is proposed that clinical symptoms manifest around Braak stage 4-5, when first limbic and then mesocortical structures become affected. Similarly, in this study the bilateral amygdalae are affected first at an early symptomatic stage, followed by reduced bilateral hippocampal volumes and thinner entorhinal, parahippocampal, and posterior cingulate regions in subsequent stages, denoting the transition to the neocortex. The simultaneous, if not earlier, implication of posterior cortical regions, however, is inconsistent with the ascending nature of pathology as proposed by Braak. This finding provides supporting evidence that multiple independent systems are affected in PD that may begin to show abnormalities at different stages of the disease.^26^

### Limitations

With respect to the HY analyses, the use of cross-sectional data does not allow us to make clear inferences on atrophy patterns and disease progression as would be the case with a longitudinal design. The HY scale also does not encompass the variety of non-motor symptoms that contribute to disease severity and progression. However, we have demonstrated that both longer illness duration and poorer cognitive performance were associated with each HY increment. Due to limited availability of medication and MoCA scores we were only able to use a subsample of the patients. Even though the MoCA subgroup was representative of the full sample in terms of demographics, there may have been hidden confounding clinical or environmental parameters influencing these results. Finally, our current multicenter analysis did not provide us with information about side-of-onset in the individual patients from the various locations. Evidence shows that disease side-of-onset is predictive for better or worse prognosis of both motor and non-motor symptoms progression.^27^ Future studies may therefore focus on how asymmetry of symptoms relates to morphological lateralisation in the brain. Follow-up analyses may include vertex-wise morphology measures to investigate localized alterations in functionally complex structures such as the thalamus and the identification of morphologically related phenotypes using unsupervised machine learning methods, in order to relate them to clinically relevant subtypes.

### Conclusions

To conclude, in this large and multi-national sample of PD patients vs. healthy controls, we found widespread structural brain abnormalities on the cortical and subcortical level that may shed new light on the pathophysiology and progression of PD. Our findings offer robust imaging signatures that are specific to the disease severity stages. The cortical and subcortical findings are strongly in line with an ongoing neurodegenerative process and with the development and extent of structural differences with increasing disease severity. The results correspond to earlier findings reported in individual studies and, importantly, also overall correspond to the staging described by Braak,^25^ with some notable exceptions. A striking finding concerns the higher thalamic volume in PD patients as compared to controls, that seems to be driven by the early stages of the disease, highlighting the need for longitudinal analyses and the inclusion of patients before the onset of motor symptoms.

## Data Availability

Data are not publicly available except for the OpenNeuro Japan, Tao Wu & Neurocon, and Parkinson's Progression Markers Initiative (PPMI) datasets.

http://fcon_1000.projects.nitrc.org/indi/retro/parkinsons.html

https://www.ppmi-info.org/access-data-specimens/

https://www.openfmri.org/dataset/ds000245/

## Acknowledgments

**ENIGMA Core**: This work was supported in part by NIH grants R56AG058854, R01AG059874, R01MH116147, R01MH117601, U54EB020403, and the Michael J Fox Foundation grant 14848, and the Kavli Foundation Neurodata without Borders

**PPMI**: Data used in the preparation of this article were obtained from the Parkinson’s Progression Markers Initiative (PPMI) database (www.ppmi-info.org/data). For up-to-date information on the study, visit www.ppmiinfo.org. PPMI – a public-private partnership – is funded by the Michael J. Fox Foundation for Parkinson’s Research and funding partners, including AbbVie, Allergan, Amathus Therapeutics, Avid Radiopharmaceuticals, Biogen, BioLegend, Bristol-Myers Squibb, Celgene, Denali, GE Healthcare, Genentech, GlaxoSmithKline, Janssen Neuroscience, Eli Lilly and Company, Lundbeck, Merck, Meso Scale Discovery (MSD), Pfizer, Piramal Enterprises, Ltd, Prevail Therapeutics, Roche, Sanofi Genzyme, Servier, Takeda, Teva, UCB, Verily and Voyager Therapeutics.

**OpenNeuro Japan**: This data was obtained from the OpenfMRI database. Its accession number is ds000245. This research was supported in part by the following: a Grant-in-Aid from the Research Committee of Central Nervous System Degenerative Diseases by the Ministry of Health, Labour, and Welfare, Integrated Research on Neuropsychiatric Disorders project, carried out by SRBPS; a Grant-in-Aid for Scientific Research on Innovative Areas (Brain Protein Aging and Dementia Control 26117002) from the MEXT of Japan; Integrated Research on Neuropsychiatric Disorders carried out under the Strategic Research Program for Brain Sciences, Scientific Research on Innovative Areas (Comprehensive Brain Science Network); and Integrated Research on Depression, Dementia, and Development Disorders by the Strategic Research Program for Brain Sciences from the Japan Agency for Medical Research and Development (AMED).

**NEUROCON**: This work was partially supported by the NEUROCON project (84/2012), financed by UEFISCDI.

**Stanford**: This work was supported by the NIH/NINDS (K23 NS075097) and the Michael J Fox Foundation for Parkinson’s Research.

**Oxford DISCOVERY**: This work was funded by Parkinson’s UK, NIHR Oxford Health Biomedical Research Centre.

**Amsterdam I:** Grant support (not for this study) from the Netherlands Organisation for Health Research and Development, the Michael J Fox Foundation and the Hersenstichting.

**Amsterdam II (Cogtips**): Cogtips is supported by the Dutch Parkinson’s Disease Association (‘Parkinson Vereniging’ 19-2015) and Brain Foundation of the Netherlands (‘Hersenstichting’ HA-2017-00227).

**Liege Cohort**: This work was funded by PDR Grant T0.165.14, National Fund for Scientific Research (FNRS), Belgium.

**Donders**: This work was funded by a grant of the Dutch Brain Foundation (grant F2013(10-15 to R.H.), from the Netherlands Organization for Scientific Research (VENI grant #91617077), and from the Michael J Fox Foundation.

**Rome SLF**: This work was funded by the Italian Ministry of Health, Italian Ministry of Health RC12-13-14-15-16-17-18-19/A.

**UNICAMP**: This work was funded by the São Paulo Research Foundation FAPESP-BRAINN (2013-07559-3).

**PDNZ**: This work was supported by the Health Research Council of New Zealand, Canterbury Medical Research Foundation, Neurological Foundation of New Zealand, University of Otago Research Grant, Brain Research New Zealand.

**Charlottesville**: This work was supported by grants from Department of Defense, Commonwealth of Virginia’s Alzheimer’s and Related Diseases Research Award Fund, and University of Virginia.

**NW-England I & II**: This work was funded by the Sydney Driscoll Neuroscience Foundation, University of Manchester Biomedical Imaging Institute, Medical Research Council UK Doctoral Training Programme, Engineering and Physical Science Research Council UK (EP/M005909/1).

**BE I & II**: Co-author for the Bern cohort receives a grant from Boston Scientific.

**UDALL & U19 Cohort**: This work was funded by the NINDS, NIH AG062418 (U19) and NS053488 (P50). We would like to acknowledge the support of Alice Chen Plotkin for MRI and clinical data collection.

**CGU**: This work was supported by MOST 106-2314-B-182-018-MY3, EMRPD1K0451, EMRPD1K0481.

**Schol-AR:** Support for this work was provided by the National Institutes of Health (grants P41EB015922 and U54EB020406) and by the Michael J. Fox Foundation’s Parkinson’s Progression Markers Initiative (PPMI – award # 8283.03).

